# The effect of age on survival in patients with peritoneal metastases from colorectal cancer who were treated with CRS-HIPEC

**DOI:** 10.1101/2025.07.28.25332334

**Authors:** Rebecca Jane Austin-Datta, Jonathan Fischer, Thomas J. George, Shantrel S. Canidate, Sophia Stanford, Faheez Mohamed, Stephen E. Kimmel

## Abstract

Discrepancies exist in the literature about the association of age and survival of patients with colorectal peritoneal metastases (CPM) who undergo cytoreductive surgery and hyperthermic intraperitoneal chemotherapy (CRS-HIPEC). We retrospectively analysed data from 811 patients with CPM aged 16 – 83 years, who were treated with CRS and HIPEC at the Peritoneal Malignancy Institute (PMI), UK, between 2000 and 2024. We ran repeated Cox proportional hazard (PH) and Kaplan-Meier survival analyses with preclinical covariates and peritoneal cancer index (PCI) covariates, to determine if associations of age with mortality hazard would be modified when the form of “age” was changed. More than 20 sets of analyses were run, each using a different form of age (continuous age, and age bins/groups e.g., <50 vs. ≥50 years). We found that within our population of pre-selected colorectal cancer patients, age at the time of CRS-HIPEC was not associated with mortality in any adjusted analyses. Early onset or late onset ages did not significantly affect survival once PCI score was included as a covariate. Discrepancies in age/survival associations reported in different studies may be due to lack of adjustment for important confounders. Our study suggests patients of any age with CPM, may benefit from CRS-HIPEC treatment.

## 2. Introduction

Peritoneal metastases (CPM) are clinically diagnosed in 5 – 15% of patients with colorectal cancer (CRC), but up to 80% of patients with CRC have been found to have colorectal peritoneal metastases (CPM) at autopsy.^1,2^ CRC diagnosis in patients aged under 50 is increasing by 2% annually.^3^ Younger patients (<50 years) are more likely to be diagnosed with advanced stage colorectal cancer (CRC) – including CPM – compared to patients older than 50 years. For example, Surveillance, Epidemiology, and End Results (SEER) data from 1998 – 2011 show younger cancer patients were more likely to be diagnosed with distant disease (metastases) than localized disease (relative risk ratio 1.58; p<0.001).^4^ Similarly, Chen et al. found that Stage III and Stage IV CRC were more likely to be diagnosed in patients under 50 years (72%) than in patients age 50 and older (63%; p=0.03).^5^ Incidence of CPM appears to increase with decreasing age: Okuno et al. report 12.5% of patients under age 40 years had CPM compared to 4.3% of patients age 40 years and older.^6^ Kaplan et al. found that for CRC patients treated in Turkey during 2003 – 2015, CPM was present in 57% patients aged 10 – 19 years and 39% patients aged 20 – 25 years, compared to just 4% of patients 26 – 94 years.^7^

Contradictory findings exist in the scarce literature on the association of age and survival of patients with CPM who receive CRS-HIPEC treatment. Studies present conflicting age-related results, reporting that compared to older patients with CPM younger patients have worse survival (e.g., Kelly et al. <45 years vs. older ages)^8^ or younger patients had better survival (e.g., Solomon et al. <50 vs. ≥50 years; Wu et al. ≤60 vs. 60 years)^9,10^, or that survival between age groups is similar (e.g., Zhou et al. ≤50 vs. >50 years).^11^ This current work set out to investigate whether such differences were attributable to the use of different age divisions. It is possible that different findings are an outcome of healthcare models or social factors related to the country of residence, rather than biological differences between different populations, or that the small sample sizes in many studies are masking associations.

Equally importantly, studies reporting associations of early/late age onset CPM with survival, do not use standard age groupings: a recent literature review found seven different methods of age division in ten CPM publications.^12^ We identified 13 different age division methods in CRC and CPM literature: seven binary methods and six ternary age groupings (Table S1). To date, no study has systematically examined the association of age with mortality in patients with CPM who received CRS-HIPEC treatment.

The goal of this study was to demonstrate how the association of survival outcomes with patient’s age (age at the time of cytoreductive surgery and hyperthermic intraperitoneal chemotherapy, CRS-HIPEC) change, when the form of age was deliberately changed in repeated (perturbed) analyses, e.g., continuous age, or different age groupings. We hypothesized that associations could differ when the form of the ‘age’ variable is changed and that the most appropriate ‘shape’ would be a U-shape spline, with younger and older patients surviving for less time; younger patients may have more aggressive cancers, older patients are more statistically likely to die based on their age.

## 3. Materials and Methods

This study analyzed data from 811 patients with CPM who were 16 – 83 years old at the time they received CRS-HIPEC treatment. Data for this project were prospectively gathered between 2000 – 2024 by the Peritoneal Malignancy Institute (PMI) in Basingstoke, United Kingdom (UK). The PMI is one of the highest-volume specialist treatment centers in the world, and part of the Hampshire Hospitals NHS (National Health Service) Foundation Trust. This study was approved as exempt by the University of Florida (UF, RRID:SCR_000145) Institutional Review Board (IRB; IRB202301936).

### Data Source and Patient Selection

Patients were drawn from a prospective database of 853 patients who had pathologically/histologically confirmed colorectal peritoneal metastases, and who had been referred to the PMI for CRS-HIPEC treatment. Patients were eligible to be included in this study if their age at time of CRS-HIPEC was known, and if post-treatment survival data were available and if their vital status (alive/dead) as of April 2024 could be confirmed in UK General Register Office records (Figure S1). These criteria were met for 811 patients. Forty-two patients in the database were removed from our analyses dataset because no post-treatment survival information was available (Figure S1).

We examined the association of post-treatment survival with CPM patient’s age at the time of CRS-HIPEC treatment (hereafter, “age”). We focused on preclinical and peritoneal cancer index (PCI) covariates. The Peritoneal Cancer Index (PCI) is a scoring system first described by Paul Sugarbaker that quantifies peritoneal disease extent and size across thirteen abdominal regions.^13^ Each region is scored from 0 to 3 based on largest lesion size in that region giving a range of scores from 0 to 39. The higher the score the more extensive the peritoneal disease; this has prognostic significance in patients undergoing CRS and HIPEC for colorectal peritoneal metastases.^8^ We included covariates in distribution analyses if at least 95% of the patients had data and if the covariate was independently associated with survival (univariate Cox PH, p≤0.2), and/or if the covariate was considered indispensable (e.g., sex). Variables in the dataset which met these requirements were: sex, treatment year, whether the patient had ever had a laparoscopy, whether the patient had their primary CRC tumor treated at the PMI at the same time as CRS-HIPEC, the American Society of Anesthesiologists physical status classification grade (ASA), if the patient was receiving neo-adjuvant chemotherapy currently (i.e., at the time of CRS-HIPEC), PCI score, and number of PCI locations with visible CPM.

Distributions of categorical variables were reported as quantity and percentages. Distributions of continuous variables were reported as median and first/third percentile (Q1, Q3). Chi-square tests were used to compare distributions of categorical variables between patients who were alive or dead. Kruskal-Wallis (K-W) tests were used to compare distributions of continuous variables between patients who were alive or dead.

Unadjusted analysis methods used were survival median (time at which 50% patients were still alive), linear regression (for continuous age only), Cox proportional hazards (Cox PH), and Kaplan-Meier (K-M) plots. Repeated (perturbed) unadjusted analyses were run, where each iteration of the analyses used a different form of the predictor variable “age”, i.e., continuous age, splines, and 17 different age groups/bins (Table 1). Age divisions and forms of age were influenced by ages reported in the CRC and CPM literature (Table S1) plus current and prior CRC routine screening ages in the United States (US) and the UK (45, 50, and 60 years).

**Table 1.**
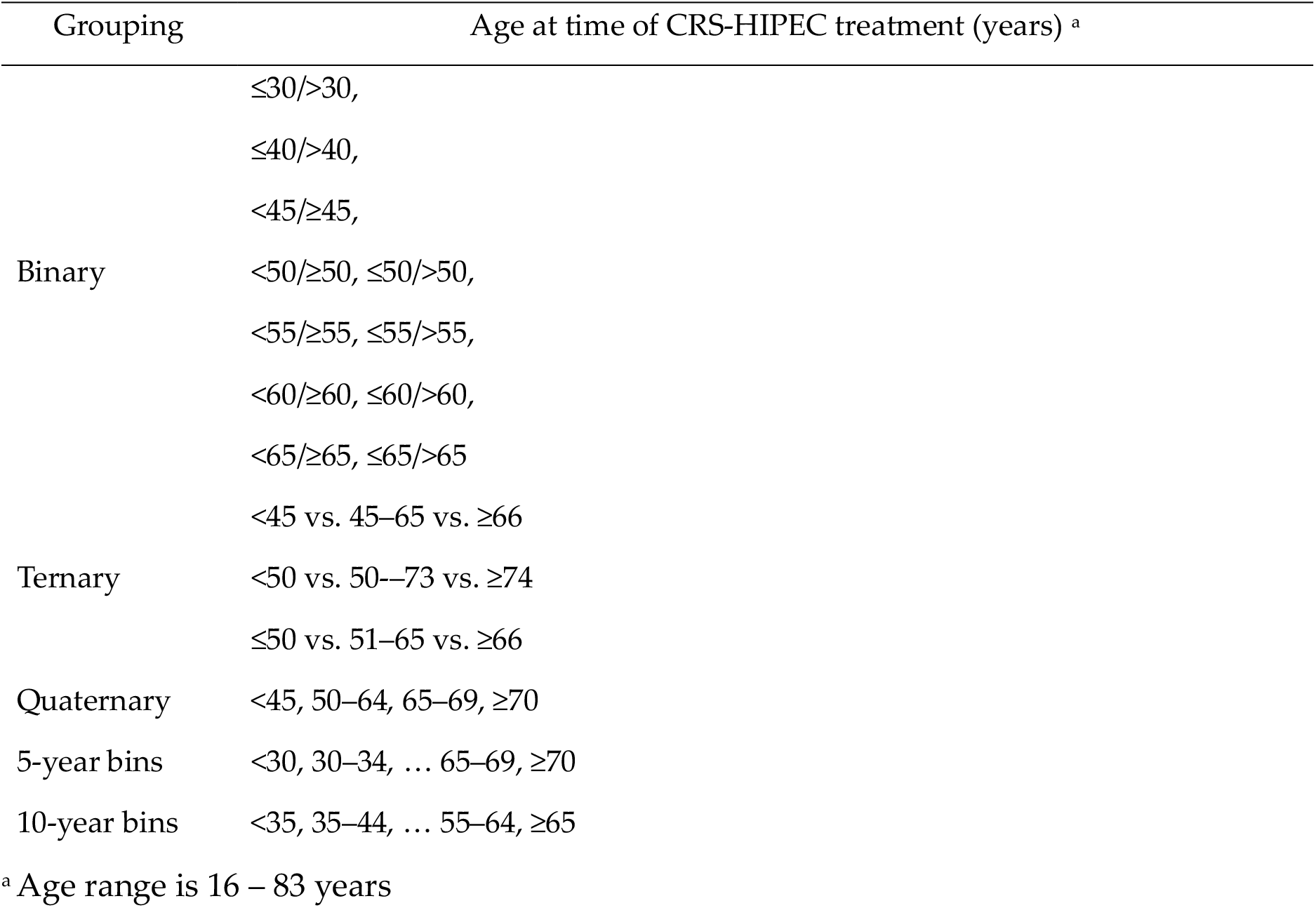
Age groups analysed in study.

Univariate Cox PH survival analyses were run for age as a continuous variable and age groups/bins. Unadjusted survival curves were plotted for age as a continuous variable and as splines. Kaplan-Meier (K-M) survival curves were plotted for continuous age and age division methods shown in Table 1. Multivariate Cox PH analyses were run for each age division method and included covariates where:

- the covariate was a pre-surgical variable, and
- a true value for the potential covariate data (i.e., not coded as ‘unknown’) was available for at least 95% of patients (i.e., ≥770 patients), and
- the univariate Cox PH survival analysis for that potential covariate had shown a p-value of p≤0.2, and
- the potential covariate had weak or no collinearity with age, via Pearson’s correlation coefficient (r-value between -0.3 and +0.3).

Covariates which met all the above criteria were: overall PCI score (range 0–39), number of locations with visible CPM (range 0–13), ASA grade (1–4), treatment year, whether the patient had ever had a laparoscopy (no/yes), whether the patient had their primary CRC tumor treated at the PMI at the same time as CRS-HIPEC (no/yes/unknown), and if the patient was receiving neo-adjuvant chemotherapy currently, i.e., at the time of CRS-HIPEC (no/yes/unknown).

Sensitivity analyses were run using a dataset of ‘complete covariates’ (n=768), which dropped patients who had an ‘unknown’ value for one or more of the selected preclinical or PCI covariates. Finally, the survival results for Binary Age Cohort B (<50, ≥50 years, patients based in the UK) were compared with survival reported by Solomon et al. for the same age division in patients with CPM who were based in the US.^9^

Statistical analyses were performed with R Software Version 4.3.3 (R Project for Statistical Computing, RRID:SCR_001905).

### Minimum detectable hazard ratio (mdHR)

Of the 811 PMI patients with post-surgery follow-up, 378 died during the study duration. There were 242 patients aged <50 years, vs. 569 patients aged 50 years or older (approximate ratio 30: 70). The minimum detectable hazard ratio (mdHR) for this binary age cohort with 811 patients was 0.73 hazard ratio, at power 80%, using two-sided alpha of 0.05, per the two-sample logrank test from https://homepage.univie.ac.at/robin.ristl/samplesize.php?test=logrank

## 4. Results

### Patient Characteristics

Patients were 16 – 83 years old at the time they underwent CRS-HIPEC (Figure S2). The median age at the time of CRS-HIPEC was 59 years (interquartile range (IQR) 48 – 68,) and 55% of patients were female (Table 2). The median PCI score recorded was 6 (IQR 3 – 11) and the median number of PCI locations was 3 (IQR 1 – 5). Primary CRC tumours were treated at the PMI for 37% patients, at other medical facilities for 60% patients; details were unknown for 3% of patients (Table 2). About half the patients had previously undergone cancer surgery (52%). Most patients had never had a laparoscopy (73%). 43 patients (6%) received neo-adjuvant chemotherapy immediately prior to CRS and HIPEC, as treatment for their primary CRC with synchronous CPM (current neo-adjuvant chemotherapy, Table 2).

**Table 2.** Characteristics of the PMI Study Population.

|  | Total<br>(N=811) | Alive<br>(N=433) | Dead<br>(N=378) | P-value |
| --- | --- | --- | --- | --- |
| **Age at time of CRS-HIPEC, years** |  |  |  | **0.019** |
| Median (Q1-Q3) | 59 (48 - 68) | 61 (49 - 68) | 57 (46 - 68) |  |
| **Gender** |  |  |  | **0.73** |
| Male | 367 (45.3%) | 193 (44.6%) | 174 (46.0%) |  |
| Female | 444 (54.7%) | 240 (55.4%) | 204 (54.0%) |  |
| **ASA Grade** |  |  |  | **0.303** |
| 1 | 56 (6.9%) | 35 (8.1%) | 21 (5.6%) |  |
| 2 | 605 (74.6%) | 316 (73.0%) | 289 (76.5%) |  |
| 3 | 133 (16.4%) | 75 (17.3%) | 58 (15.3%) |  |
| 4 | 6 (0.7%) | 2 (0.5%) | 4 (1.1%) |  |
| Missing | 11 (1.4%) | 5 (1.2%) | 6 (1.6%) |  |
| **PCI Score (0-39)** |  |  |  | **<0.001** |
| median (Q1-Q3) | 6.0 (3.0 - 11) | 5.0 (3.0-6.0) | 9.0 (5.0 - 15) |  |
| **Visible CPM, number of locations** |  |  |  | **<0.001** |
| median (Q1-Q3) | 3.0 (1.0-5.0) | 2.0 (1.0-3.0) | 4.0 (2.0 - 7.0) |  |
| **Was primary CRC tumour treated at the PMI?** |  |  |  | **0.002** |
| No | 488 (60.2%) | 261 (60.3%) | 227 (60.1%) |  |
| Yes | 301 (37.1%) | 151 (34.9%) | 150 (39.7%) |  |
| Unknown | 22 (2.7%) | 21 (4.8%) | 1 (0.3%) |  |
| **Previous Surgery for Cancer?** |  |  |  | **0.0743** |
| No | 379 (46.7%) | 214 (49.4%) | 165 (43.7%) |  |
| Yes | 418 (51.5%) | 209 (48.3%) | 209 (55.3%) |  |
| Unknown | 14 (1.7%) | 10 (2.3%) | 4 (1.1%) |  |
| **Laparoscopy, ever?** |  |  |  | **<0.001** |
| No | 588 (72.5%) | 350 (80.8%) | 238 (63.0%) |  |
| Yes | 223 (27.5%) | 83 (19.2%) | 140 (37.0%) |  |
| **Current neo-adjuvant chemo?** |  |  |  | **0.281** |
| No | 762 (94.0%) | 404 (93.3%) | 358 (94.7%) |  |
| Yes | 43 (5.3%) | 27 (6.2%) | 16 (4.2%) |  |
| Unknown | 6 (0.7%) | 2 (0.5%) | 4 (1.1%) |  |

### Mortality

When the study concluded in mid-April 2024, 378 patients (47%) had died (Table 2). The survival median after CRS-HIPEC treatment for all patients (accounting for censoring) was 48 months (95% confidence interval (CI) 43, 57; Figure S3). Patients who died during the study period tended to be younger at the time of CRS-HIPEC treatment (median age 57 years; IQR 46 – 68) compared to patients who were censored (median age 61 years; IQR 49 – 68; p=0.019, Table 2). Patients who died also had higher PCI (median PCI 9 for deceased, vs. 5 for censored patients; p<0.001, Table 2), had more locations with visible CPM (median 4 PCI locations for deceased, vs. 2 for censored patients; p<0.001, Table 2), and were less likely to have undergone a laparoscopy procedure (19% of deceased had ever had a laparoscopy vs. 37% of censored patients; p<0.001, Table 2). Patient sex, ASA grade, primary CRC tumour treatment at the PMI, previous surgery for cancer, and current neo-adjuvant chemotherapy were not significantly different comparing patients who died or who were censored (Table 2).

A summary of survival results (survival medians, and uni-and multivariate Cox PH HRs) for continuous age, current/prior country-specific ages recommended to initiate CRC screening, and a ternary age group cohort is shown in Table 3. In each age grouping, although univariate Cox PH HR may have shown significant differences between groups, the survival median 95% CI ranges overlapped between groups (i.e., no clinically relevant difference). This was confirmed when the related K-M plots showed overlapping confidence interval bands (Figures 1, S4, and S5). Subsequently, any significance between age groups that we observed in univariate analyses, was lost in multivariate analyses. Brief descriptions are below; more details including results from 5-year and 10-year age bins are in Supplemental Materials.

**Table 3.**
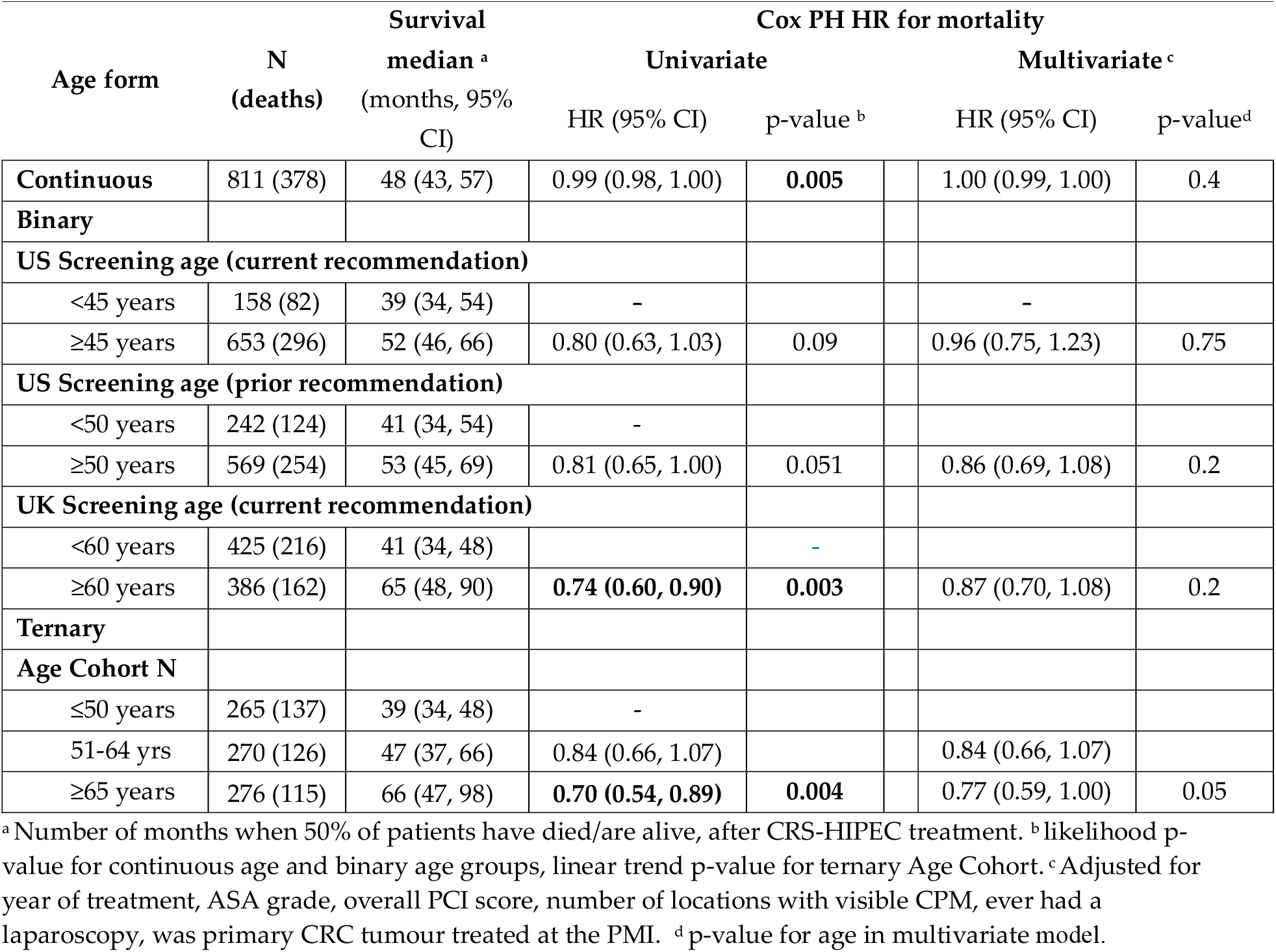
Examples of Univariate and Multivariate Cox PH Outcomes for continuous age, and selected binary/ternary age groups.

**Figure 1.**
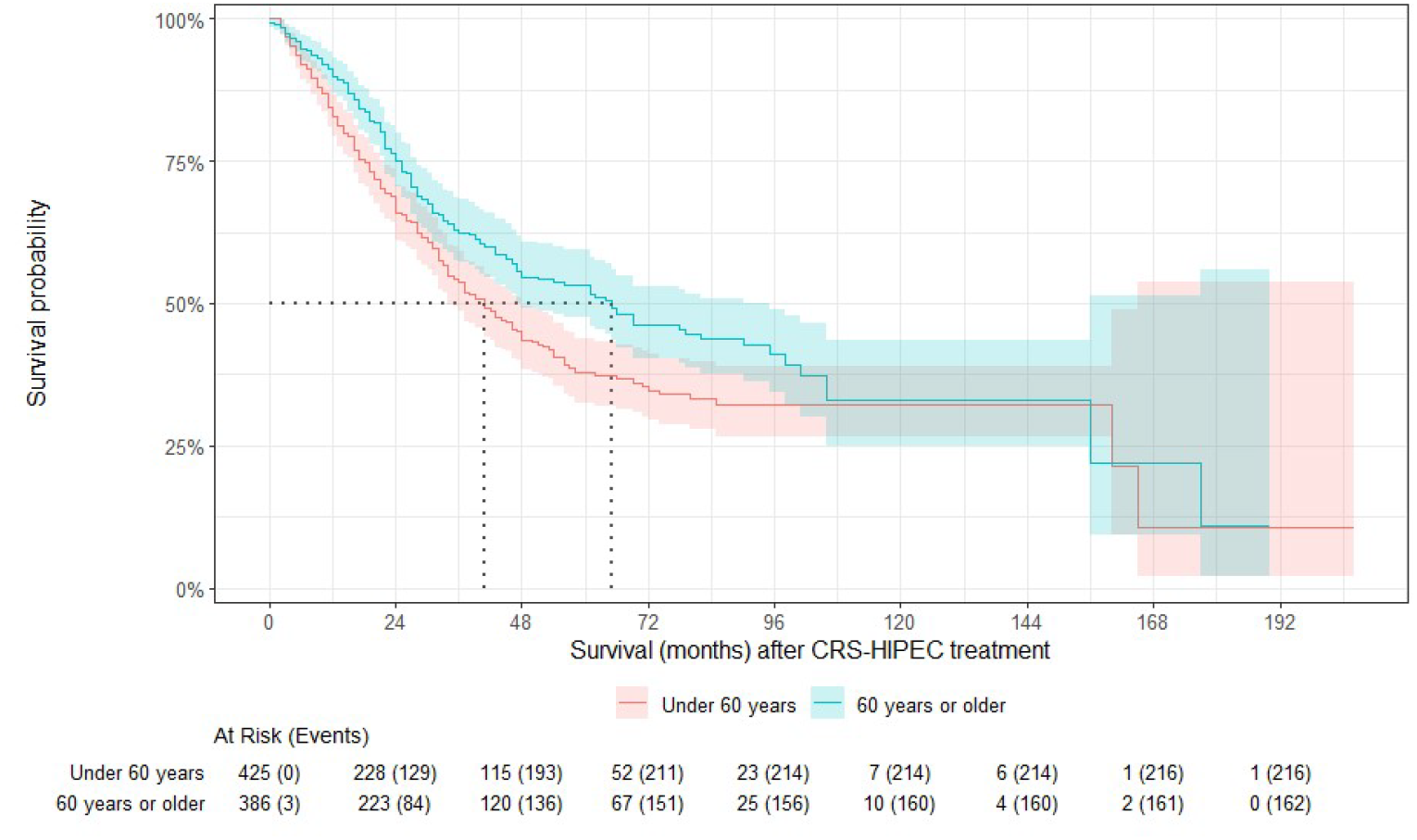
K-M plot <60 vs. ≥60 years (Age Cohort F; current UK CRC screening age cut-off); shows survival medians and 95% CI bands.

### Continuous age - Univariate and multivariate analyses

In unadjusted survival analyses using continuous age, for every one-year increase in patient age (at the time of CRS-HIPEC), the patient would be expected to survive an additional 0.19 months (p=0.03).

However, after adjustment, the multivariate Cox PH HR for continuous age became “1.00”, i.e., no association between survival and patient age at time of CRS-HIPEC (95% CI 0.99, 1.00; p=0.4, Table 3).

### Splines

During exploration of the role age may play in survival after CRS-HIPEC treatment, several spline options were tested for ‘survival ∼ age’ using unadjusted Cox PH methods. For example, P-spline with 1, 2, 3, and 4 degrees of freedom, B-Splines with 3, 4, 13, and 50 degrees of freedom and Cox PH spline. There was no indication of an age-survival relationship for the PMI patients using splines (Figures 2, S6, S7, and S8).

**Figure 2.**
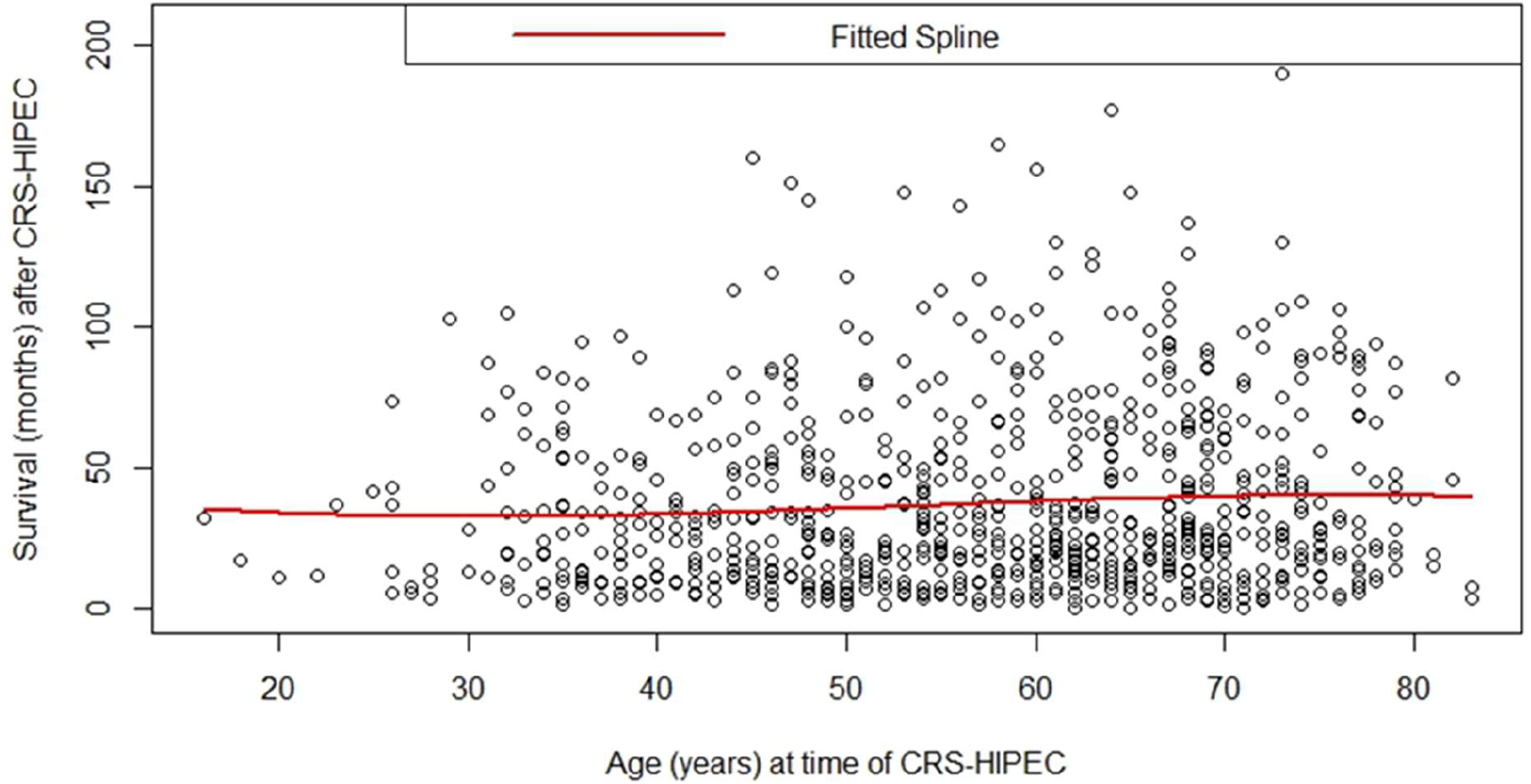
B-spline regression (3 degrees freedom), survival by age at time of CRS-HIPEC.

### Binary age cohorts – Univariate and multivariate analyses

Patients were divided into binary age cohorts using nine age division methods inspired by methods used in CRC and CPM literature and cut-off ages for current and prior screening ages in the UK and the US: ≤40/>40,^6,14^ <45/≥45 (US current screening age), <50/≥50 (US prior screening age),^9^ ≤50/>50,^11^ <55/≥55, ≤55/>55, <60/≥60 (UK current screening age),^15^ ≤60/>60, <65/≥65,^16,17^ and ≤65/>65 years.^8^ For completeness, we also ran *post hoc* analyses for ≤30/>30 years (See supplemental materials). In each binary age cohort, the older group had a greater survival median value (accounting for censoring), compared to the younger group (Tables 3 and S3). However, in all binary Age Cohort K-M plots, the 95% confidence interval bands for survival overlapped (Figures 1, S4, S5, and S9 through S16).

The univariate Cox PH hazard ratio for mortality was between 0.72 and 0.81 for each binary age cohort, where the younger group was used as the reference, i.e. the older age group had lower hazard of mortality (Table S3). After adjustment for covariates, Cox PH HR for binary age cohorts shifted towards the null; none of the multivariate hazard ratios were statistically significant for age. However, all the HRs remained below 1.0. For example, the univariate Cox PH hazard ratio for <60/≥60 years was 0.74 (95% CI 0.60, 0.90; p=0.003) but significance was lost in multivariate Cox PH analysis (HR 0.87; 95% CI 0.70, 1.08; p=0.21, Table 3).

### Ternary/quaternary age cohorts – Univariate and multivariate analyses

Patients were divided into five different ternary/quaternary age cohorts using age division methods inspired by methods used in CRC literature: <45/45 – 65/≥66,^8^ <50/50 – 73/≥74,^18^ <50/50 – 64/65 – 69/≥70,^19^ ≤50/51 – 64/≥65, ≤50/51 – 65/≥66 years.

The univariate Cox PH HR for three of the ternary Age Cohorts (K, N, P) had a univariate statistically significant p-value for trend, with the oldest group in each of these cohorts (≥66, ≥65, and ≥66 years respectively) having significantly lower mortality hazard compared to the youngest group (<45 or ≤50 years, Table S4). In all four of the ternary Age Cohorts, the survival median (accounting for censoring) increased from youngest to oldest age group (Table S4). In Quaternary Age Cohort M (<50/50 – 64/65 – 69/≥70 years) patients aged 65-69 years had the longest survival median (95 months) compared to all other age groups (all age division methods, accounting for censoring), but insufficient patient numbers/events in this age group prevented generation of the survival’s upper 95% CI, so this finding should be interpreted with caution (Table S4). Regardless, in multivariate Cox PH models, Cox PH HR for ternary age cohorts shifted towards the null, and none of the results were statistically significant.

We tested each individual variable used in the Cox PH multivariate model for continuous age (Table S2) in an “age + covariate” bivariate model to see which individual variables were driving the loss of statistical significance for age. Only two of the variables led to a noticeable change in age-specific HR, in the bivariate model: PCI score (Age HR=0.996, 95% CI 0.989, 1.003; p=0.3), and number of locations with visible CPM (Age HR=0.997, 95% CI 0.989, 1.004; p=0.4); these changes were not statistically significant.

## 5. Discussion

We examined associations of survival with age as: (a) continuous age (regression, splines), (b) age as a binary/ternary/quaternary variable, and (c) in modified 5- and 10-year categorical age groups. In adjusted Cox PH models, none of the age forms used (continuous, splines, or age division methods) indicated that age is associated with multivariate adjusted survival with statistical significance in our population.

PCI score remained significantly associated with mortality in the multivariate model (p<0.001). This finding mirrors conclusions from an expert consensus group, who determined that PCI is a major – and perhaps the most important – variable associated with survival of patients with CPM.^2^ However, PCI scores are rarely reported in analyses. It is possible that the lack of PCI reporting reflects a lack of available data: e.g., although PCI may be reported in HIPEC studies, PCI may not be included in general CRC datasets that were not originally intended to incorporate CRS-HIPEC. If researchers are unable to use PCI score in their analyses, and if PCI is the most significant mortality predictor, then age-survival associations that are not adjusted for PCI could be considered flawed.

We expected to see that binary age divisions would provide very different results depending on which ages were included in the lower and upper age groups. (e.g., different hazard ratios, with p-values <0.05). We thought that younger aged patients would survive less time than patients of median age in the dataset– because literature infers that cancers in younger patients appear to be more aggressive than older adults’ cancers. We thought that older patients - based on their age – would be statistically more likely to die than median-aged patients.

We showed that different forms of age will give slightly different results in univariate and multivariate Cox PH analyses (Tables 3, S3, S4, S5, and S6). Binary age divisions gave different survival results depending on which ages were included in the lower and upper age groups (Table S3). Some univariate Cox PH age-survival results were statistically significant at p<0.05. However, once covariates were included in age-survival analyses (multivariate Cox PH), age-mortality Cox PH ratios for all forms of age became statistically insignificant (Tables S3, S4, S5, and S6).

To support generalizability of the data, we compared age-survival results for patients with CPM who received CRS-HIPEC at the PMI (Basingstoke, United Kingdom), with those who received CRS-HIPEC at Mount Sinai Hospital (MSH, New York City, United States).^9^ The K-M survival plots for patients with CPM who were <50 vs. ≥50 years, showed mirror-image curves for the older groups treated at PMI vs. MSH (Figure S5 in this work vs. Figure 1b in the MSH paper).^9^

Survival for younger patients was very similar between the two treatment centres (<50 years PMI: survival median 41 months; <50 years MSH: median follow-up 42 months - survival median not reached). Differences observed were driven by survival of the PMI and MSH ‘older patients’: survival median for patients aged ≥50 years at PMI was 53 months, almost double the survival median for ≥50 years treated at MSH (28 ’months, p=0.011).^9^ Age was significantly associated with survival in multivariate analyses for MSH patients (HR 0.43, 95% CI 0.21, 0.91; p=0.026), but not for PMI patients (HR 0.86, 95% CI 0.69, 1.08; p=0.2).

Sensitivity analyses were run to see if the opposite survival for older MSH and PMI patients could be explained by treatment era or a diagnosis/procedure time lag. However, none of the sensitivity analyses changed the overall direction of PMI results: in each case PMI older patients had greater survival than younger patients.

When comparing the MSH and PMI survival data, we should bear in mind the potential for differing risks of referral bias, including potential referral bias related to concerns that treatment may result in financial toxicity (FT) for an underinsured patient in the United States (MSH).^20^ For PMI patients, the cost of CRS-HIPEC treatment and hospital stay would be covered by the National Health Service (UK). This healthcare system difference may limit some extrapolation of the results to the US population. Regardless, patients with CPM who are not referred for or not treated with CRS-HIPEC would likely be those deemed to have CPM too extensive to benefit from treatment, further contributing to referral and selection bias in our dataset.

Alternatively, patients with CPM who were not referred for treatment may have had comorbidities that were not conducive to procedure under anaesthesia. Comorbidities may be more likely in older patients, potentially creating a false association between older age at the time of CRS-HIPEC and greater survival.

The cross-sectional and vital status patient data used in this study were provided by a specialist medical institute (the PMI), who focused on collecting data related to each patient’s current CPM diagnosis, and the CRS-HIPEC treatment performed by their location. ‘Survival’ is the only longitudinal variable to which we have access. Due to the PMI’s focused data collection, we have no information on potentially informative sociodemographic variables e.g. income, education level, or lifestyle factors (e.g., body mass index, diet, smoking status), or family medical history. Due to patient privacy concerns, we were not provided with information on race/ethnicity. Another limitation of our data includes the lack of a comparison group. We only have data for patients with CPM who did receive CRS-HIPEC treatment. We do not have survival information for patients with CPM who did not receive treatment.

Limitations notwithstanding, this study is, to our knowledge, the first reported ‘perturbed’ analyses of mortality hazard using different forms of age (e.g., continuous age and different age groups) for patients with CPM. Having granular PCI information was possibly the greatest strength of the PMI dataset. Adjustment for PCI score resulted in a non-significant association between age and mortality.

The PMI patients received CRS-HIPEC treatment for CPM, during a 25-year period (2000–2024), providing a robust long-term set of outcomes data. All 811 patients in our analyses have vital status confirmed; patients who did not originally have a date of death recorded in the dataset were confirmed as alive or dead by cross-referencing the PMI data in death records held by the UK General Register Office. It is uncommon to have confirmed vital status available for patients in studies covering a 25-year period.

## 6. Conclusion

It seems statistically likely that patients with CPM of any age would benefit equally from CRS-HIPEC treatment. The results of our adjusted analyses indicate that in our population of patients with CPM, age is not significantly associated with survival after CRS-HIPEC treatment. However, Cox PH mortality hazards did alter when different age forms or groupings were used; this showed that standard age cut-points would facilitate direct comparisons between studies and make meta-analyses a practical option. We recommend that the methods used in our analyses are repeated in other populations.

## Data Availability

All data produced in the present work are contained in the manuscript and supplemental materials (available on medRxiv). The analyses used hospital data and thus the raw data are not openly available to protect patient privacy. De-identified datasets may be requested from the Peritoneal Malignancy Institute. The R code used to run analyses are available in Supplemental Material.

## Acknowledgments

We would like to acknowledge Camran Nesari for assistance with data in the early stages of this project. We would also like to acknowledge the staff and team at the Peritoneal Malignancy Institute.

## Ethical Statement

The study’s protocol (which uses de-identified hospital data, i.e., data without personal identifier variables), was reviewed and approved as exempt by the University of Florida (UF) Institutional Review Board (IRB; IRB202301936).

## Data Accessibility

The article is based on hospital data and is thus not openly available to protect patient privacy. De-identified datasets may be requested from the Peritoneal Malignancy Institute. The R code used to run analyses are available in Supplemental Material.

## Declaration of AI use

We have not used AI-assisted technologies in creating this article.

## Authors’ Contributions

RJAD: conceptualization, data curation, formal analysis, investigation, methodology, project administration, resources, software, supervision, validation, visualization, writing – original draft, writing – review & editing; JF: conceptualization, formal analysis, methodology, software, supervision, validation, visualization, writing – review & editing; TJG: conceptualization, formal analysis, methodology, supervision, writing – review & editing; SSC: conceptualization, methodology, writing – review & editing; SS: data curation, formal analysis, investigation, project administration, resources, software, writing – review & editing; FM: conceptualization, data curation, investigation, project administration, resources, writing – review & editing; SEK: conceptualization, formal analysis, methodology, project administration, supervision, visualization, writing – original draft, writing – review & editing.

