## Supplementary material for "The effect of age on survival in patients with peritoneal metastases from colorectal cancer who were treated with CRS-HIPEC": R Code

**R code used for analyses for article:** The effect of age on survival in patients with peritoneal metastases from colorectal cancer who were treated with CRS-HIPEC. Austin-Datta et al. 2025-07-28

```
########################
# R CODE FOR PAPER BY AUSTIN-DATTA ET AL. 2025-07-28
# TITLE: The effect of age on survival in patients with peritoneal metastases from colorectal cancer
# who were treated with CRS-HIPEC
########################
# LOAD LIBRARIES
########################
library(survival) # Needed to run Cox PH
library(summarytools) # Create frequency tables
library(gtsummary) # Survival median (get n, events, survival value with 95% CIs)
library(ggsurvfit) # Create time-to-event figures
library(splines) # Create spline figures
library(table1) # Create Table 1 (population characteristics)
library("Hmisc") # Adds labels for table1 code (NOTE: Summary tools masks Hmisc)
########################
# SET DATA PATHS
########################
# Set working directory as folder where the dataset is housed
setwd("") # insert file path between the "" marks
# Read in the dataset, copy the dataset and work with the copy
# If dataset already loaded, suggest reloading to avoid issues with variable
formats
df100 <- read.csv("DataFileName.csv")
df32 <- df100 # Copy in case of snafus
########################
# FORMAT VARIABLES AS REQUIRED (e.g., numeric, or factored)
########################
# Set the required format for the variables
# Sample codes to put variables in numeric and factored formats are shown below.
# Our multivar Cox PH analyses were adjusted by four variables in numeric format
# (year of treatment, ASA grade, overall PCI score, locations with visible CRC-PM)
# and two variables factored Yes=1, No=0 (laparoscopy; PMI primary tumour
treatment).
 # NOTE: For Cox PH, K-M plots, survival median calculations, numeric format
needed
# for vital status (e.g., Dead1RtCens0) and survival duration (e.g.,
SurvivalMonths)
```

```
# BUT Table1 code requires vital status to be factored
########################
# NUMERIC FORMAT - CODE
########################
# Examples of forcing variables into numeric format:
df32$Dead1RtCens0 <- as.numeric(df32$Dead1RtCens0) # Vital status, numeric
df32$SurvivalMonths <- as.numeric(df32$SurvivalMonths) # Survival duration, 
numeric
 ########################
# FACTORED FORMAT - CODE
########################
# Factor and label categorical variables using base R,
# including variables based on age group sections:
# For example, the age division "Under 45 years at time of CRS-HIPEC"
# has variable name U45y1n0
# it is coded in the dataset as 1=Under 45 years, 0=45 years and older
df32$U45y1n0 <- factor(df32$U45y1n0, levels=c(1,0) # (1=U45, 0=45+)
  labels=c("<45 years","≥45 years"))
########################
# BASE-R FREQUENCY TABLE CODE
########################
freq(df32$U45y1n0) # Run frequency table to check n are as expected
########################
# SURVIVAL PACKAGE - COX PH UNIVARIATE
########################
# Example of how to run univariate Cox PH for "Under 45 years at time of CRS-
HIPEC"
# using factored U45y1n0, and numeric vars SurvivalMonths and Dead1RtCens0
# library(survival) # Run univariate and multivariate Cox PH, using Survival 
package
CoxModU45 <- coxph(Surv(SurvivalMonths, Dead1RtCens0) ~ U45y1n0
                  , data=df32)  # build univariate model
summary(CoxModU45)  # model summary
########################
# SURVIVAL PACKAGE - COX PH MULTIVARIATE
########################
CoxModMultU45 <- coxph(Surv(SurvivalMonths, Dead1RtCens0) ~ U45y1n0
```

```
      + NumericYear
      + NumericASAgrade
      + FactoredPCI_Score_Surg # Overall PCI score at time of surgery
      + FactoredN_PCI_locs # Number of PCI locations with visible CPM
      + FactoredLapY1N0 # Did patient have a laparoscopy ever yes/no
      + FactoredPTumY1N0 # Primary CRC tumor treated at PMI yes/no
                  , data=df32)  # build model on full data
summary(CoxModMultU45)  # model summary
########################
# SURVIVAL MEDIAN WITH 95% CONFIDENCE INTERVALS (SURVIVAL with GTSUMMARY)
########################
# Example: how to get survival median value for "Under 45 years at time of CRS-
HIPEC"
# Using the factored variable U45y1n0
# library(gtsummary) # Survival median (n, events, survival median with 95% CIs)
survfit(Surv(SurvivalMonths, Dead1RtCens0) ~ U45y1n0, data = df32)
######################
# KAPLAN-MEIER PLOTS WITH 95% CONFIDENCE INTERVALS
########################
# Kaplan-Meier plots showing survival median values, and a risk table underneath
# Example of how to get K-M plot for "Under 45 years at time of CRS-HIPEC"
# Using the factored variable U45y1n0
# library(ggsurvfit) # Plot K-M curves
# Kaplan-Meier plots by Binary Age U45 v 45-and-older
p <- survfit2(Surv(SurvivalMonths, Dead1RtCens0) ~ U45y1n0, data = df32) %>%
        ggsurvfit() +
        scale_ggsurvfit() + # Change Y axis into percentage
        scale_x_continuous(limits = c(0, 206),
                     breaks =
                       c(0,24,48,72,96,120,144,168,192)) + # 24-month intervals
        add_quantile(y_value = 0.5,
                    linetype = "dotted",
                    color = "grey30", linewidth = 0.8) + # Median dotted lines
        add_confidence_interval() +
        labs(title = "Kaplan-Meier plot, <45 v. ≥45 years",
                x = "Survival (months) after CRS-HIPEC treatment",
                y = "Survival probability"
```

```
  ) + add_risktable(risktable_stats = "{n.risk} ({cum.event})")
p
p2 <- p + add_censor_mark() # Adds censoring marks
p2
########################
# SPLINE CODE – Example for B-spline with 3 degrees of freedpom
########################
# Recommend you reload the dataset to avoid issues with variable formats
df1 <- df100 # Reload the dataset and call it a new name
# library(splines)
spline_model <- lm(df1$SurvivalMonths ~ bs(df1$AgeSurgYears, df = 3))
# Here, ns from the splines package is used to create a B-spline basis with 3
degrees of freedom.
# Visualize the data and fitted spline
plot(df1$AgeSurgYears, df1$SurvivalMonths,
     main = "B-Spline Regression Example with 3df",
     xlab = "df1$AgeSurgYears",
     ylab = "df1$SurvivalMonths")
lines(df1$AgeSurgYears, predict(spline_model), col = "red", lwd = 2)
legend("topright", legend = "Fitted Spline", col = "red", lwd = 2)
########################
# TABLE ONE CODE
########################
# Recommend you reload the dataset to avoid issues with variable formats
df2 <- df100 # Reload the dataset and call it a new name
########################
# Load the packages for Table 1
library(table1) # This is required to run Table 1 code
library("Hmisc") # Needed to add labels for Table 1
# Note: when writing code put Hmisc 'label' code *underneath* factoring code
########################
# NOTE: EXPORTING TABLE 1 WHEN DONE
# To copy-paste Table1 Output into word doc,
# Copy table using cmd+a -> cmd+c -> paste Special in MS Word -> chose .html
format
########################
# Set the required format for the variables (e.g., numeric, factored.)
```

```
# This Table 1 distribution analysis uses categorical + numeric formats variables
# Sample codes to put variables in numeric and factored formats, shown below
########################
# FACTOR, THEN ADD HMISC LABLE
########################
# NOTE: For Table 1 make sure that vital status (i.e., Dead1RtCens0) is factored
# Note: put Hmisc variable 'label()' code *underneath* the factoring code
# Run ‘freq’ code to double-check data are factored properly
df2$Dead1RtCens0 <-
  factor(df2$Dead1RtCens0,
         levels=c(0,1),
         labels=c("Alive", # Censored
                  "Dead"))
label(df2$Dead1RtCens0) <- "Vital Status in April 2024" # Hmisc label for Table1
freq(df2$Dead1RtCens0) # Double-check data are factored properly
########################
# FACTOR AND LABEL CATEGORICAL VARIABLES
########################
# For example, ‘sex’
df2$SexM1F2 <-
  factor(df2$SexM1F2,
         levels=c(1,2),
         labels=c("Male", # Reference
                  "Female"))
label(df2$SexM1F2) <- "Sex" # Hmisc label is used in Table1
freq(df2$SexM1F2) # Double-check data are factored properly
########################
# FACTOR AND LABEL VARIABLES BASED ON AGE GROUPS
########################
# For example, the age division “Under 45 years at time of CRS-HIPEC”
# has variable name U45y1n0 (1=Under 45 years, 0=45 years and older)
df2$U45y1n0 <-
  factor(df2$U45y1n0, levels=c(1,0),  # (1=U45, 0=45 years and older)
         labels=c("<45 years","≥45 years"))
label(df2$U45y1n0) <- "Binary Age, <45 v. ≥45 years" # Hmisc label is used in
Table1
# Note: put Hmisc variable 'label()' code *underneath* the factoring code
```

```
########################
# NUMERIC FORMAT
########################
# Examples of forcing variables into numeric format:
df2$Age = as.numeric(df2$Age)
df2$PCI_Score_Surg = as.numeric(df2$PCI_Score_Surg) # PCI score at time of surgery
df2$N_PCI_locs = as.numeric(df2$N_PCI_locs) # Number of PCI locations with CPM
########################
# Write Hmisc label codes *underneath* the “as.numeric” code (otherwise R may crash):
label(df2$Age) <- "Age at time of CRS-HIPEC" # This is continuous age
label(df2$PCI_Score_Surg) <- "PCI Score (0-39)"
label(df2$N_PCI_locs) <- "Number of locations with visible PCI"
########################
# UNIT LABELLING GOES LAST
########################
units(df2$Age) <- "years"
########################
# To prepare for Table1 code, make a ‘RENDER’ function
# Note: Code below is based on work from
# https://benjaminrich.github.io/table1/vignettes/table1-examples.html
########################
# Code to switch in Median with Q1, Q3
my.render.cont <- function(x) {
  with(stats.apply.rounding(stats.default(x, ), digits = 2),
       c("",
         "median (Q1-Q3)" =
           sprintf(paste("%s (",Q1,"- %s)"), MEDIAN,Q3)))
}
########################
# Use ‘RENDER’ function in first run of Table 1 code
# First run will give column percentages (total on the left), and median/Q1-Q3 values
########################
#library(table1)
table1(~ Age + SexM1F2+ ASA + PCI_Score_Surg+ N_PCI_locs
       + PrimaryTumour + PrevSurg +Lap + CurrentNeoAdjChemoId
```

```
       | Dead1RtCens0, data=df2,
overall=c(left="Total"),render.continuous=my.render.cont)

# This will give you the column total percentages on the left, and median/Q1-Q3
values

########################

# Prepare to add p-value column in second run of Table 1 code

# Based on https://benjaminrich.github.io/table1/vignettes/table1-examples.html

########################

# Create a p-value function

# Add in p-value columns

pvalue <- function(x, ...) {

  # Construct vectors of data y, and groups (strata) g

  y <- unlist(x)

  g <- factor(rep(1:length(x), times=sapply(x, length)))

  if (is.numeric(y)) {

    # For numeric variables, perform a standard 2-sample t-test

    p <- t.test(y ~ g)$p.value

  } else {

    # For categorical variables, perform a chi-squared test of independence

    p <- chisq.test(table(y, g))$p.value

  }

  # Format the p-value, using an HTML entity for the less-than sign.

  # The initial empty string places the output on the line below the variable
label.

  c("", sub("<", "&lt;", format.pval(p, digits=3, eps=0.001)))

}

########################

# Use ‘p-value’ and ‘RENDER’ functions in second run of Table 1 code

# Second run gives p-values (on right) plus column percentages median/Q1-Q3 values

########################

#library(table1)

table1(~ Age + SexM1F2+ ASA + PCI_Score_Surg+ N_PCI_locs

       + PrimaryTumour + PrevSurg +Lap + CurrentNeoAdjChemoId

       | Dead1RtCens0, data=df2, overall=F,

       render.continuous=my.render.cont, extra.col=list(`P-value`=pvalue))

########################

# https://cran.r-project.org/web/packages/table1/vignettes/table1-examples.html
```

```
# https://benjaminrich.github.io/table1/vignettes/table1-examples.html
```

### Data accessibility

Dataset generated from patient records, dataset is not open access to preserve patient privacy.

### Declaration of AI use

AI-assisted technologies were not used to write the R code above
