## Supplemental Materials for "The effect of age on survival in patients with peritoneal metastases from colorectal cancer who were treated with CRS-HIPEC"

### Details of Preliminary Work

#### Determining Covariates to be used in Multivariate Cox PH

**Univariate Cox PH survival for preclinical/PCI covariates – 95% complete data**

We ran univariate Cox PH survival analyses for all preclinical/PCI variables with data for 95% or more patients (birth year, sex, treatment year centered on 2015, overall PCI score, number of PCI locations where CPM was visible, ASA grade, whether the patient had previously undergone cancer surgery and/or had their primary CRC tumor treated at the PMI and/or had ever had a laparoscopy, and/or had previous adjuvant or current/previous neo-adjuvant chemotherapy).

Eight covariates with data for ≥95% patients, were independently associated with survival (univariate Cox PH p≤0.2, Table S2). These potential confounders were:

- overall PCI score (p<2e-16)
- number of PCI locations where CPM was visible (p<2e-16)
- treatment year centered on 2015 (p=2.15e-07)
- laparoscopy procedure, ever (no/yes, p=0.0001)
- primary CRC tumor treated at PMI (no/unknown/yes, p=0.02)
- ASA grade (p=0.049)
- birth year (p=0.1)
- current neo-adjuvant chemotherapy (no/unknown/yes, p=0.2).

**Univariate Cox PH survival – 'Unknown' removed from individual variables**

Any continuous preclinical/PCI variables with a missing/unknown value were automatically excluded from univariate Cox PH calculations, e.g., ASA grade (treated as 'continuous' variable) was missing for 11 patients, so the R output showed n=800 (rather than n=811 per the dataset used for analysis). However, five of the categorical variables had 'unknown' included as a category (primary CRC tumor treated at the PMI, previous cancer surgery, previous adjuvant or current/previous neo-adjuvant chemotherapy).

To ensure that 'unknown' status was not distorting univariate Cox PH hazard ratios, we ran univariate Cox PH analyses with only those patients who had yes/no responses for the five variables, i.e., we excluded 'unknown' response from the analysis. We also ran univariate Cox PH analyses for overall PCI score, with 'zero' values removed (data shown in dissertation). In some cases, when categorical variables were confined to only yes/no responses per the 'complete dataset', their univariate Cox PH analysis association with survival changed (data shown in dissertation). For example:

- previous adjuvant chemotherapy (no/yes, p=0.4; vs. no/unk/yes, p=0.5)
- previous neo-adjuvant chemotherapy (no/yes, p=0.2; vs. no/unk/yes, p=0.4)
- current neo-adjuvant chemotherapy (no/yes, p=0.1; vs. no/unk/yes, p=0.2)
- primary CRC tumor treated (no/yes, p=0.5; vs. no/unk/yes, p=0.02)

The likelihood ratio p-value for PCI score (p<2e-16) did not change whether 'zero' values were included or excluded in univariate Cox PH analysis. Similarly, the likelihood ratio p-value for previous cancer surgery (p=0.8) did not change whether patients with 'unknown' were included or excluded.

**Univariate Cox PH survival – sensitivity analyses with 100% complete data**

A 'complete' dataset was created by removing patients who had 'unknown' for one or more of five specific categorical covariates (primary CRC tumor, previous cancer surgery, previous adjuvant or current/previous neo-adjuvant chemotherapy).

There were 768 patients in the 'complete dataset', each of whom had a known value for all of the five categorical covariates listed above. The 'complete dataset' was used to run a set of univariate Cox PH sensitivity analyses for the 768 patients, using the preclinical/PCI variables we had used in regular univariate analyses (birth year, sex, treatment year centered on 2015, overall PCI score, number of PCI locations, ASA grade, previous cancer surgery, primary CRC tumor, laparoscopy, previous adjuvant or current/previous neo-adjuvant chemotherapy). The p-value for 'Primary tumor treated at PMI' lost significance (p=0.45) while having a record of being prescribed neo-adjuvant chemotherapy (previously, or currently) both became significant (p=0.16 for both; full table shown in dissertation).

**Association of Age with Non-Age Preclinical/PCI Covariates**

Pearson's correlation analyses were run for selected preclinical/PCI numeric variables to determine collinearity with age (Pearson's product-moment correlation >0.75). Birth year and age had correlation of -0.96, "age" was retained in the multivariate Cox PH model; other numeric variables were not correlated with age.

To check if age at time of CRS-HIPEC was associated with any non-age preclinical/PCI covariates, we ran regular univariate regression for the preclinical/PCI variables noted above, using the overall data and also for only those patients who had yes/no responses for categorical variables and those who had non-zero overall PCI score, i.e., we excluded 'unknown' or 'zero PCI' response from analyses.

**Selecting Cox PH Model for Analyses – Original Dataset (n=811)**

First, we ran four different multivariate Cox PH analyses with continuous age on the original dataset (n=811). The models differed in the pre-surgical/PCI covariates that were used. We selected covariates independently associated with survival (Cox PH HR univariate analysis) with significance of either p≤0.2, or p≤0.05. For each level of univariate significance, we ran a multivariate model with the covariates where at least 70% (568/811) or at least 95% of patients (711/811) had data, i.e.,

- Model A used variables with univariate Cox PH significance of p≤0.2 where ≥70% of patients had data,
- Model B used variables with univariate Cox PH significance of p≤0.2 where ≥95% of patients had data,
- Model C used variables with univariate Cox PH significance of p≤0.05 where ≥70% of patients had data, and
- Model D used variables with univariate Cox PH significance of p≤0.05 where ≥95% of patients had data.

The main difference as regards covariates included in each multivariate model, was whether ECOG, tumor grade, tumor histology and/or ASA were included in each model, based on their univariate Cox PH p-values and percent complete data. For example, ECOG, tumor grade and histology all had univariate Cox PH of p≤0.05, and ≥70% complete data, while ASA had p≤0.2 but ≥95% complete data. Correlations between pairs of variables were checked: ECOG and ASA had no relationship (Pearson's correlation, r=0.12, i.e., r<0.25), tumor grade and tumor histology also had no relationship (Pearson's correlation, r= 0.23, i.e., r<0.25). There were no real differences in results between the four multivariate models as regards the Cox PH hazard ratio for 'age'.

Model B (p≤0.2, 95% complete data) was selected as the final 'original dataset' multivariate model to be reported (Table S2). The covariates used in the final 'original dataset' multivariate model were overall PCI score, number of locations with visible CPM, treatment year, whether the patient had ever had a laparoscopy, whether the patient had their primary CRC tumor treated at the PMI at the same time as CRS-HIPEC, ASA grade, and whether the patient was currently receiving neo-adjuvant chemotherapy (Table S2). Birth year was highly correlated with Age at CRS-HIPEC, so birth year was excluded from the final multivariate model.

#### Additional Information about Methods

**Covariate Selection**

We focused on covariates which could potentially be used in pre-treatment clinical decision-making tools (e.g., nomograms to determine viability of CRS-HIPEC treatment), including covariates related to the severity and extent of CPM (e.g., PCI score). We excluded surgical and post-operative variables (e.g., *length of CRS-HIPEC procedure and post-operative complications) because they could not be used in pre-treatment clinical decision-making tools.* We replaced a set of 13 binary variables "PCI location X has visible CPM" (Yes/No), with a single numeric variable 'Number of PCI locations with visible CPM' which recorded how many of the 13 PCI location areas had visible evidence of CPM.

**Non-normal distribution**

Each of the continuous numeric variables used had non-normal distribution, e.g., age at time of CRS-HIPEC treatment. This was determined by running four different tests of normality (skew, kurtosis, Jarque-Bera Normality Test, Kolmogorov-Smirnov (K-S) test) and examining box-and-whisker plots and Q-Q plots**.**

When the PMI dataset was initiated in 2000, CRS-HIPEC treatment was relatively novel and only a few patients were treated each year in the early 2000's (Figure S23). Most patients in the PMI dataset were treated after 2015 – so patients surviving treatment in earlier years were skewing the survival estimates i.e., creating lead-time bias. This issue was partly addressed by centering treatment year on 2015 in analyses, but non-normal distribution in this variable remained. Hence, in distribution analyses we presented the median and inter quartile range (Table 2).

**Statistical Analyses**

Statistical analyses were run using R Software Version 4.3.3, 2024-02-29 ucrt (R Project for Statistical Computing, RRID:SCR_001905). The R packages used include base R (RRID:SCR_001905),[1] '_table1' to create demographic tables (RRID:SCR_024900),[2] 'survival' to run Cox PH analysis including "survfit" from the survival package to calculate survival medians (RRID:SCR_021137).[3] To create and format hazard ratio tables, 'gtsummary' including 'gt' from the gtsummary package was used (RRID:SCR_021319).[4] To create labels for tables, R packages 'summarytools' (RRID:SCR_026818)[5] and 'Hmisc' (Harrell Miscellaneous RRID:SCR_022497) were used.[6] To create plots, we used ggplot2 (RRID:SCR_014601)[8,] and ggsurvfit (RRID:SCR_025045).[7,8] R codes used in this article, with related notes, are available in the electronic supplemental material.

#### Additional Information about Results

**K-M Plots by age**

K-M plots comparing survival by age group show overlapping 95% CI bands, which indicate a lack of statistical difference between the groups:

- Binary division: Figures 1, S4, S5, and S10 through S16
- Ternary/Quaternary division: Figures S17 through S22,
- 5-year age bins: Figures S24 and S25
- 10-year age bins: Figures S26 and S27.

**Summary of Age Comparison Results**

Patients who died tended to be younger at the time of CRS-HIPEC treatment (median age 57 years; IQR 46-68) and patients who survived tended to be older at the time of treatment (median age 61 years; IQR 49-68). This difference was statistically significant (p=0.019, Table 2).

The youngest binary age group (≤30 years at time of CRS-HIPEC) had the lowest survival median (28 months) of any age group analyzed, with univariate Cox PH hazard ratio 0.46 for ≤30/>30 years (95% CI 0.27, 0.78; p=0.004).

When patients were divided into age groups the survival median and univariate Cox PH survival trends were not statistically significant for age cohorts that included dividing ages 70 or 74 years (p>0.05; Ternary Age Cohort L (Table S4), Quaternary Age Cohort M (Table S4); 10-year age bins Cohort R in Table S6). Once covariates were included in multivariate Cox PH models, age was no longer statistically significant (Tables S4 and S6).

**Binary age cohorts – details of univariate survival results**

**Age Cohort A, ≤40/>40 years:** The younger and older groups in Age Cohort A (≤40 vs. >40 years) had survival median of 37 and 50 months respectively, with univariate Cox PH HR of 0.78 (95% CI 0.59, 1.03) but this was not significant (p=0.076; Table S3). The K-M survival curve for Age Cohort A shows that after the first three months, better survival was experienced by patients who were older than 40 at the time of CRS-HIPEC, compared to younger patients. This advantage continued until 156 months when the survival curves crossed; however, at this time there was only one patient in the younger group (Figure S10); at 156 months the real-time age division for the "≤40/>40 years" cohort was ≤53/>53 years.

**Age Cohort B, <50/≥50**: The younger and older groups in Age Cohort B (<50 vs. ≥50 years) had survival median of 41 and 53 months respectively, with univariate Cox PH HR of 0.81 (95% CI 0.65, 1.0) but this also was not significant (p=0.051; Table S3). The K-M survival curve for Age Cohort B shows that after the first 12 months, better survival was experienced by patients who were 50 or older at the time of CRS-HIPEC, compared to younger patients. This advantage continued until 156 months when the survival curves crossed (Figure S5). At 156 months, the current age division for the "<50/≥50 years" cohort would be <63/≥63 years, when there were less than ten patients in each group.

**Age Cohort C, ≤50/>50:** The younger and older groups in Age Cohort C (≤50 vs. >50 years) had survival median of 39 and 56 months respectively, with a statistically significant univariate Cox PH HR of 0.76 (95% CI 0.62, 0.64; p=0.012; Table S3). The K-M survival curve for Age Cohort C shows that after the first three months, better survival was experienced by patients who were older than 50 years at the time of CRS-HIPEC, compared to younger patients. This advantage continued to 156 months when the survival curves crossed (Figure S11). At 156 months, the current age division for the "≤50/>50 years" cohort would be ≤63/>63 years, at which time six or fewer patients were still alive in each group.

**Age Cohort D, <55/≥55:** The younger and older groups in Age Cohort D (<55 vs. ≥55 years) had survival median of 41 and 62 months respectively, with a statistically significant univariate Cox PH HR of 0.73 (95% CI 0.60, 0.90; p=0.003; Table S3). The K-M survival curve for Age Cohort D shows that after the first three months, better survival was experienced by patients who were 55 or older at the time of CRS-HIPEC, compared to younger patients. This advantage continued until 156 months when the survival curves crossed (Figure S12). At 156 months, the current age division for the "<55/≥55 years" cohort would be <68/≥68 years, with five patients or fewer alive in each group.

**Age Cohort E, ≤55/>55:** The younger and older groups in Age Cohort E (≤55 vs. >55 years) had survival median of 41 and 65 months respectively, with a statistically significant univariate Cox PH HR of 0.73 (95% CI 0.59, 0.89; p=0.002; Table S3). The K-M survival curve for Age Cohort E shows that after the first three months, better survival was experienced by patients who were older than 55 at the time of CRS-HIPEC, compared to younger patients. This advantage continued until 156 months when survival curves crossed (Figure S13). At 156 months, the current age division for the "≤55/>55 years" cohort would be ≤68/>68 years, with five patients or fewer alive in each group.

**Age Cohort F, <60/≥60:** The younger and older groups in Age Cohort F (<60 vs. ≥60 years) had survival median of 41 and 65 months respectively, with a statistically significant univariate Cox PH HR of 0.74 (95% CI 0.60, 0.90; p=0.003; Table S3). The K-M survival curve for Age Cohort F shows that after the first three months, better survival was experienced by patients who were 60 or older at the time of CRS-HIPEC, compared to younger patients. This advantage continued until 108 months when the survival curves tracked very similar survival (Figure 1). At 108 months, the current age division for the "<60/≥60 years" cohort would be <69/≥69 years, and there were 25 patients or fewer still alive in each group.

**Age Cohort G, ≤60/>60:** The younger and older groups in Age Cohort G (≤60 vs. >60 years) had survival median of 41 and 65 months respectively (Table S3). The univariate Cox PH HR was 0.72 (95% CI 0.58, 0.88; p=0.002; Table S3). This was the lowest HR of all binary models, i.e., this ≤60 vs. >60 years age division showed the greatest difference in mortality hazard for all binary ages tested. The K-M survival curve for Age Cohort G shows that after the first three months, better survival was experienced by patients who were older than 60 at the time of CRS-HIPEC, compared to younger patients. This advantage continued until about 109 months when survival curves merged (Figure S14). At 109 months, the current age division for the "≤60/>60 years" cohort would be ≤69/>69 years, at which time 25 patients or fewer were alive in each group.

**Age Cohort H, <65/≥65:** The younger and older groups in Age Cohort H (≤65 vs. >65 years) had survival median of 43 and 66 months respectively, with a statistically significant univariate Cox PH HR of 0.76 (95% CI 0.61, 0.95; p=0.014; Table S3). The K-M survival curve for Age Cohort H shows that after the first three months, better survival was experienced by patients who were 65 or older at the time of CRS-HIPEC, compared to younger patients. This advantage continued until 108 months when the survival curves tracked very similar survival (Figure S15). At 108 months, the current age division for the "<65/≥65 years" cohort would be <74/≥74 years, at which time there were about 32 "younger" and 16 "older" patients alive in each group.

**Age Cohort J, ≤65/>65:** The greatest difference in survival median between younger and older groups, (23 months difference) was for patients aged 65 years or less vs. patients who were older than 65 years at the time of CRS-HIPEC (43 months vs. 66 months, Cox PH HR 0.76; p=0.016, Age Cohort J, Table S3). The K-M survival curve for Age Cohort J shows that after the first three months, better survival was experienced by patients who were older than 65 at the time of CRS-HIPEC, compared to younger patients. This advantage continued until about 102 months when survival curves crossed (Figure S16). At 102 months, the current age division for the "≤65/>65 years" cohort would be ≤73.5/>73.5 years, at which time there were about 34 "younger" and 14 "older" patients alive in each group.

**Post hoc analysis, ≤30/>30:** For completeness, an additional analysis was run for patients divided into ≤30/>30 years (prompted by the 5-year age group results, discussed elsewhere): the univariate Cox PH hazard ratio for ≤30/>30 years was 0.46 (95% CI 0.27, 0.78; p=0.004) which indicated patients older than 30 years at the time of CRS-HIPEC treatment had about half the (unadjusted) hazard of dying compared to patients aged 30 years or younger. The younger group had the lowest survival median of any age group analyzed (≤30 years, survival median 28 months; 95% CI 13, 43; Figure S9) while the older group (>30 years) had survival median of 48 months (95% CI 44, 61, Figure S9). (Note: these data were not included in Table S3 as this sub-analysis was not part of the original analysis plan). The K-M survival curve for the post hoc binary cohort shows that after the first 12 months, better survival was experienced by patients who were older than 30 at the time of CRS-HIPEC, compared to younger patients. This advantage continued until about 102 months, at which time all the patients in the younger group had died (Figure S9).

**Age-Survival Univariate Analyses – Trend for grouped ages**

When patients were divided into age groups there was a trend for patients who were ***older*** (but not necessarily the oldest) at the time of CRS-HIPEC to have a ***greater*** survival median (time at when 50% of participants were still alive) compared to the youngest patient group (p<0.05), for binary, ternary/quaternary, and sets of modified ten-year/five-year age bins. This was true for:

- binary groups with patients aged >50 years in the older group (p<0.05)
- ternary groups' patients ≥65 years vs. <45, or vs. ≤50 years (p<0.05)
- 10-year age bin 60-69 years vs. patients <40 years (p<0.01)
- 5-year age bin 55-64 and ≥65 years, vs. patients <35 years (p<0.05)
- 5-year age bin 55>60 and older, vs. patients <30 years (p<0.05).

This trend is shown in survival median columns of Tables S3, S4, S5, and S6).

The quaternary cohort's group of patients aged 65-69 years appeared to have the longest survival (95 months) compared to younger or older groups, but insufficient events in this age group prevented generation of the survival's upper 95% confidence interval, so this finding may be interpreted with caution (Table S4).

The quaternary group adds an upper age category onto a ternary age division method used by Siegel et al.[9]

**Modified 5-year age bins – Univariate and multivariate analyses**

Patients were divided into modified 5-year age bins: <30, 30>35, 35>40, 40>45, 45>50, 50>55, 55>60, 60>65, 65>70, 70>75, and ≥75 (Table S5). K-M plots showed substantially overlapping 95% CI bands for the 5-year age bins (Figures S24 and S25).

We ran survival median calculations (accounting for censoring) for each age bin: while there was an overall trend for older patients (up to age 70) to have greater survival median compared to younger patients, the trend was absent between ages 35-55 (Table S5). The <30-year age bin had the lowest survival median of any age bin in any age division in our analyses (32 months, Table S5). Due to relatively low numbers of patients in each 5-year age bin it was not possible to calculate upper 95% CIs for survival median values in seven of the nine age bins, hence survival median values may be unreliable (Table S5).

The univariate Cox PH analysis for 5-year age bins had a non-significant linear trend p-value (p=0.065, Table S5). Cox PH HRs indicated lower hazard for 5-year age bins 55>60 and older, compared to age group <30. For example, age bin 55>60 had HR of 0.50 (95% CI 0.27, 0.95; Table S5). There were no significant differences for mortality hazards between the five-year age bins in multivariate analysis (HR linear trend p=0.057; Table S5).

**Modified 5-year age groups – details of survival results**

We had intended to run survival analyses in strict 5-year age groups. However, there were small numbers of patients in the 5-year groups of youngest and oldest ages (Table S5). The jitterplots of mean survival for youngest groups were similar: Figure S28 shows the mean error bar for each of the three younger age groups include the mean values of the other two younger age groups. Also, the mean survival values of the oldest groups were similar, and the 75->80 age group mean error bars are completely within the 80->85 error bar range. Similar overlap occurs for vital status of these age groups (Figure S29). Therefore, we decided to combine the three youngest and two oldest age groups into categories 'Younger than 30 years' and '75 years and older' (Table S5).

Survival median for patients younger than 30 years was only 32 months. When survival of 5-year age groups was compared to survival of patients <30 years, there were significant hazard ratios for age groups 55 years and older. The maximum survival median of 95 months was in the 65>70-year age group (HR 0.34; 95% CI 0.18, 0.65; Table S5). However, these results must be interpreted with caution because the relatively low number of events (deaths) in the modified "5-year" age groups prevented generation of values for an upper 95% confidence interval for survival time in nine of the eleven age bins (Table S5). Although in the modified 5-year age bin Cox PH survival analyses patients under age 35 years had the worst mean survival compared to older groups, these results were inconclusive due to relatively low numbers of patients in each 5-year age bin. Neither univariate nor multivariate Cox PH HRs for 5-year age bins were statistically significant (Table S5).

**Modified 10-year Age Bins – Univariate and Multivariate Analyses**

We ran univariate analyses in two sets of modified 10-year age groups, with <35 and <40 years as the youngest age groups (Age Cohorts Q and R). In 10-year Age Cohort Q, the youngest age group (<35 years) had the lowest survival median of any group in our analyses (34 months, Table S6). Univariate Cox PH hazard ratios showed lower hazard ratio for older age groups up to age 70, with statistically significant linear trend p-values of 0.006 and 0.033 for Age Cohorts Q and R respectively (Table S6). However, K-M plots showed overlapping confidence intervals, indicating lack of real significant differences (Figure S26, S27). Additionally, multivariate Cox PH analyses were not significant (Table S6).

**Lack of race/ethnicity covariates in dataset due to privacy concerns**

The UK NHS has mandated since 1995 that patient's ethnicity must be recorded when patients are admitted to an NHS hospital.[10] The PMI is part of the NHS. However, limited numbers of CPM patients were treated each year during 2000-2024, especially in the earlier years. Unique combinations of age, ethnicity, vital status, and treatment date for each patient could be used to identify individuals. Therefore, to protect patient privacy, ethnicity was not included in the dataset that the PMI provided for this project.

We know the 811 PMI patients included in this study all had a UK 'National Insurance Number (NIN)', i.e., they were drawn from a population of UK residents. Race/ethnicity information in the UK census periods 2011 and 2021 show vast majority of UK residents self-identify as White (86%, 81.7%).[11] Basingstoke, where the PMI is located, is in Southern England; it is reasonable to infer the majority of referred patients with NIN would be from England/Wales rather than Scotland (patients in the north of England and Scotland would be expected to be referred to The Christie NHS Foundation Trust Oncology Center in Manchester, https://www.christie.nhs.uk ). In the UK 2021 census, self-identified ethnicity demographics of the respondents from England/Wales were 81.7% White, 9.3% Asian, 2.5% Black or African; these numbers record fewer people self-identifying as "White" and more people identifying as Asian, Black, or African compared to the UK census ten years earlier.[11] The 2011 census recorded race/ethnicity self-identification data as 86.0% White, 7.5% Asian, 1.8% Black or African.[11] A study by Delon et al. (2022) reported CRC incidence in England during 2013–2017 in similar proportions of ethnicity as the 2011 census. Over 32 thousand CRC cases were reported; CRC patients were 90% White, 2.1% Asian, 1.4% Black.[12]

Based on the UK census data (majority of people in the UK self-identifying as White in 2011 and 2021) and based on the CRC incidence 2013-2017 in England, we could infer that most CPM patients referred to the PMI may self-identify as White, rather than Black, African, or Asian. However, we cannot be certain of this, as the English CRC patient data were for all CRC cancer stages, not specific to CPM.[12]

**Consideration of variables' association with survival**

When the study concluded in April 2024, 433 patients (53%) were known to be alive, while 378 patients (47%) were known to have died. Median follow-up time was 35 months for patients who were censored (i.e., still alive) at the end of the study (IRQ 14-69), while survival median was 22 months (IQR 11-36) for patients who had died (p=<0.001). The gender distribution for alive and dead patients mirrored the 45%/55% male/female distribution observed overall (Table 2).

Overall, 52% (418/811) of the patients had previously had surgery for cancer, but 55% of patients who died and 48% of patients who were alive in April 2024, had previously had surgery for cancer but this difference was not statistically significant (p=0.0743, Table 2). It was more common for patients to be asymptomatic rather than symptomatic at the time of their initial oncologic presentation (24% vs. 35%) but symptomatic status was frequently unknown (40%, Table 2). Patients who were alive at the end of the study were more likely to have been asymptomatic (42%) compared to patients who died (42% vs. 26%; p<0.001; Table 2).

Most patients never had a laparoscopy (73%). Patients who died were more likely to have 'ever' had a laparoscopy compared to patients who were alive at the end of the study (63% vs. 81%; p<0.001; Table 2). Most patients were not receiving neo-adjuvant chemotherapy at the time of CRS-HIPEC (94%), and there was no statistically significant difference between patients who were alive compared to those who died (93% vs. 95%; p=0.281; Table 2). Most patients presented with ASA grade 2 or 3, i.e. 'Relevant disease' (75%) or 'Restrictive disease' (16%); no significant different was shown between those who had died or were alive at the end of the study (Table 2).

PCI was significantly associated with survival (p<0.001; Table 2). The PCI overall scores recorded for patients ranged from 0-39, with median PCI score of 6 (IQR 3-11). Patients who died during the study had median PCI of 9 (IQR 5-15) while patients who were alive at the end of the study had median PCI of 5 (IQR 3-6). The number of locations in the peritoneum where PCI was visible, was also significantly associated with survival (p<0.001, Table 2). The number of PCI locations recorded for patients ranged from 0-13, with median of 3 locations (IQR 1-5). Patients who died during the study had a median of 4 PCI with visible CPM recorded (IQR 2-7) while patients alive at the end of the study had a median of 2 locations with visible CPM recorded (IQR 1-3).

Table S1. Binary/ternary age groupings and patient numbers used in CRC/CPM literature.

| **Reference** | **Young group** | **n (%)** | **Middle group** | **n (%)** | **Older group** | **n (%)** |
|---|---|---|---|---|---|---|
| Okuno et al.[13] | <40 | 57 (10) | *NA* | *NA* | ≥40 | 513 (90) |
| Haleshappa et al.[14] | <40 | 89 (88) | *NA* | *NA* | 40 | 11 (12) |
| Solomon et al.[15] | <50 | 43 (44) | *NA* | *NA* | ≥50 | 45 (56) |
| Zhou et al.[16] | ≤50 | 46 (33) | *NA* | *NA* | >50 | 94 (67) |
| Péron et al.[17] | <60 | 422 (53) | *NA* | *NA* | ≥60 | 373 (47) |
| Moraes Filho et al.[18] | <65 | NR* | *NA* | *NA* | ≥65 | NR* |
| Xiao et al.[19] | <65 | 14 (45) | *NA* | *NA* | ≥65 | 17 (55) |
| Kelly et al.[20] | <45 | 202 (18) | 45-65 | 549 (48) | >65 | 387 (34) |
| Seigel et al.[9] | <50 | 19,550 (13)[a] | 50-64 | 48,210 (32)[a] | 65>70 | 85,260 (56)[a] |
| Ulanja et al.[21] | 18-49 | RC 2881 (7)[b]<br>LC 5537 (15)[b] | 50-64 | RC 10,366 (27)[b]<br>LC 14,567 (40)[b] | ≥65 | RC 25,124 (66)[b]<br>LC 16,229 (45)[b] |
| Lurvink et al.[22] | <50 | SPM 27 (7)[c]<br>MPM 24 (7)[c] | 50->74 | SPM 249 (61)[c]<br>MPM 221 (68)[c] | ≥74 | SPM 133 (33)[c]<br>MPM 81 (25)[c] |
| Asghari-Jafarabadi et al.[23] | <50 | RA 74 (5)[d]<br>LC 145 (14)[d] | 50-75 | RA 669 (46)[d]<br>LC 552 (54)[d] | >75 | RA 710 (49)[d]<br>LC 325 (32)[d] |
| Fataftah et al.[24] | <60 | 39 (39) | 60-70 | 33 (33) | >70 | 28 (28) |

[a] Estimated US CRC 2023 data; [b] 2005-2019 SEER data; [c] percentage calculated from table data; Abbreviations: LC, left colon (splenic flexure, descending colon, sigmoid, rectosigmoid); MPM, metachronous peritoneal metastases (diagnosed >90 days post CRC surgery); NR* Not reported (main analytical focus was right/left side cancer; numbers of patients in binary age groups used to calculate hazard ratio for older/younger patients were not reported); RC, right colon (cecum, ascending colon, hepatic flexure, transverse colon); RA, right colon/appendix (appendix, caecum, ascending colon, hepatic flexure, transverse colon); LC, left colon (splenic flexure, descending colon, sigmoid colon, rectosigmoid); SPM, synchronous peritoneal metastases (diagnosed before or ≤90 days post CRC surgery).

Table S2. Univariate and Multivariate Cox PH Hazard Ratios for mortality for Continuous Age and Pre-surgical and PCI Variables for Full Dataset (n=811, events=378)

| | | **Univariate Cox PH for mortality** N=811, events = 378 | | | Multivariate **Cox PH for mortality** [c] N=811, events = 378 | |
|---|---|---|---|---|---|---|
| **Variable** | **Range/cats** | **HR (95% CI)** | **p-value** | **Range/cats** | **HR (95% CI)** | **p-value** |
| Age (Continuous) | 16.8-83.0 | 0.99 (0.98, 1.00) | **0.005** | 16.8-83.0 | 1.00 (0.99, 1.00) | 0.4 |
| Birth year [a] | 1925-1998 | 1.01 (1.00, 1.01) | 0.11 | - | - | - |
| Treatment Year[b] | 2000-2024 | 0.93 (0.90, 0.96) | **<0.001** | 2000-2024 | 0.96 (0.94, 0.99) | **0.020** |
| ASA grade (n=800) | 1-4 | 1.22 (1.00, 1.49) | **0.050** | 1-4 | 1.21 (0.99, 1.48) | 0.069 |
| PCI score | 0-39 | 1.09 (1.08, 1.10) | **<0.001** | 0-39 | 1.06 (1.03, 1.09) | **<0.001** |
| # Visible CPM locations | 0-13 | 1.21 (1.18, 1.24) | **<0.001** | 0-13 | 1.07 (1.00, 1.15) | 0.051 |
| Sex | Male<br>Female | -<br>0.95 (0.77, 1.16) | 0.61 | - | - | - |
| Laparoscopy | No<br>Yes | -<br>1.50 (1.22, 1.86) | **<0.001** | No<br>Yes | 1.32 (1.06, 1.64) | **0.014** |
| Previous cancer surgery | No<br>Yes<br>Unknown | -<br>1.02 (0.83, 1.25)<br>0.73 (0.27, 1.98) | 0.78 | - | - | - |
| Primary CRC tumor treated at PMI | No<br>Yes<br>Unknown | -<br>1.07 (0.87, 1.32)<br>0.15 (0.02, 1.06) | **0.016** | No<br>Yes<br>Unknown | -<br>1.01 (0.81, 1.25)<br>0.24 (0.03, 1.73) | 0.2 |
| Previous adjuvant chemotherapy | No<br>Yes<br>Unknown | -<br>1.10 (0.90, 1.35)<br>0.62 (0.15, 2.50) | 0.47 | - | - | - |
| Previous neo-adjuvant chemotherapy | No<br>Yes<br>Unknown | -<br>1.20 (0.93, 1.54)<br>0.89 (0.22, 3.58) | 0.38 | - | - | - |
| Current neo-adjuvant chemotherapy | No<br>Yes<br>Unknown | -<br>1.60 (0.97, 2.64)<br>1.18 (0.44, 3.17) | 0.22 | - | - | - |

[a] Birth year was highly correlated with Age at CRS-HIPEC, so was dropped from the multivariate model. [b] Centered on 2015. [c] Adjusted for year of treatment, ASA grade, overall PCI score, number of locations with visible CPM, ever had a laparoscopy, was primary CRC tumor treated at the PMI. Definitions: “Current neo-adjuvant chemotherapy” - neo-adjuvant chemotherapy given immediately prior to CRS and HIPEC, as treatment for CPM with or without the primary colorectal cancer in place. “Previous neo-adjuvant chemotherapy” – systemic therapy given prior to surgical resection of the primary colorectal cancer. “Previous adjuvant chemotherapy” – systemic therapy given after surgical resection of the primary colorectal tumour.

Table S3. Binary age cohorts: Survival Median and Cox PH Hazard Ratios for mortality.

| | n (deaths) | Survival median [a] | Univariate Cox PH | | Multivariate Cox PH [b] | |
|---|---|---|---|---|---|---|
| | | (months, 95% CI) | HR (95% CI) | p-value | HR (95% CI) | p-value |
| **Age Cohort A** | | | | | | |
| ≤40 years | 113 (61) | 36.9 (32, 54) | - | - | | |
| >40 years | 698 (317) | 49.9 (45, 64) | 0.78 (0.59, 1.03) | 0.076 | 0.96 (0.72, 1.27) | 0.77 |
| **Age Cohort B** | | | | | | |
| <50 years | 242 (124) | 41 (34, 54) | - | - | | |
| ≥50 years | 569 (254) | 53 (45, 69) | 0.81 (0.65, 1.00) | 0.051 | 0.86 (0.69, 1.08) | 0.19 |
| **Age Cohort C** | | | | | | |
| ≤50 years | 265 (137) | 39 (34, 48) | - | - | | |
| >50 years | 546 (241) | 56 (46, 69) | **0.76 (0.62, 0.94)** | **0.012** | 0.81 (0.65, 1.00) | 0.05 |
| **Age Cohort D** | | | | | | |
| <55 years | 327 (169) | 41 (34, 48) | - | - | | |
| ≥55 years | 484 (209) | 62 (47, 79) | **0.73 (0.60, 0.90)** | **0.003** | **0.78 (0.63, 0.96)** | **0.02** |
| **Age Cohort E** | | | | | | |
| ≤55 years | 346 (179) | 41 (34, 47) | - | - | | |
| >55 years | 465 (199) | 65 (48, 82) | **0.73 (0.59, 0.89)** | **0.002** | **0.79 (0.64, 0.97)** | **0.02** |
| **Age Cohort F** | | | | | | |
| <60 years | 425 (216) | 41 (34, 48) | | - | | |
| ≥60 years | 386 (162) | 65 (48, 90) | **0.74 (0.60, 0.90)** | **0.003** | 0.87 (0.70, 1.08) | 0.21 |
| **Age Cohort G** | | | | | | |
| ≤60 years | 448 (228) | 41 (34, 48) | | - | | |
| >60 years | 363 (150) | 65 (48, 95) | **0.72 (0.58, 0.88)** | **0.002** | 0.83 (0.67, 1.03) | 0.10 |
| **Age Cohort H** | | | | | | |
| <65 years | 535 (263) | 43 (37, 52) | | - | | |
| ≥65 years | 276 (115) | 66 (47, 98) | **0.76 (0.61, 0.95)** | **0.014** | 0.84 (0.67, 1.06) | 0.14 |
| **Age Cohort J** | | | | | | |
| ≤65 years | 556 (271) | 43 (37, 52) | | - | | |
| >65 years | 255 (107) | 66 (47, 98) | **0.76 (0.61, 0.95)** | **0.016** | 0.85 (0.67, 1.07) | 0.16 |

[a] Number of months when 50% of patients have died/are alive, after CRS-HIPEC treatment. [b] Adjusted for year of treatment, ASA grade, overall PCI score, number of locations with visible CPM, ever had a laparoscopy, was primary CRC tumor treated at the PMI.

Table S4. Survival median and Univariate Cox PH Hazard Ratios for mortality (Qua/Ternary age cohorts).

| | **n (deaths)** | **Survival median** [a] | **Univariate Cox PH for mortality** | | **Multivariate Cox PH for mortality** [d] | |
|---|---|---|---|---|---|---|
| | | (months, 95% CI) | **HR** (95% CI) | **p-value** [c] | **HR** (95% CI) | **p-value** [c] |
| **All patients** | 811 (378) | 48 (43, 57) | 0.99 (0.98, 1.00) | 0.004 | 1.00 (0.99, 1.00) | 0.4 |
| **Age Cohort K** | | | | **0.013** | | 0.32 |
| <45 years | 158 (82) | 39 (34, 54) | - | - | | |
| 45-65 years | 398 (189) | 47 (37, 56) | 0.88 (0.68, 1.14) | - | 1.02 (0.78, 1.33) | |
| ≥66 years | 255 (107) | 66 (47, 98) | **0.69 (0.52, 0.93)** | - | 0.86 (0.64, 1.16) | |
| **Age Cohort L** | | | | 0.22 | | 0.91 |
| <50 years | 242 (124) | 41 (34, 54) | - | - | | |
| 50-73 years | 490 (218) | 53 (45, 69) | 0.81 (0.65, 1.01) | - | 0.85 (0.68; 1.06) | |
| ≥74 years | 79 (36) | 56 (38, NA) [b] | 0.79 (0.55, 1.15) | - | 1.02 (0.69, 1.51) | |
| **Age Cohort M** | | | | 0.22 | | 0.86 |
| <50 years | 242 (124) | 41 (34, 54) | - | | | |
| 50-64 years | 293 (139) | 46 (35, 62) | 0.90 (0.71, 1.15) | - | 0.92 (0.72, 1.17) | |
| 65-69 Years | 125 (45) | 95 (51, NA) [b] | **0.59 (0.42, 0.83)** | - | **0.61 (0.43, 0.86)** | |
| ≥70 years | 151 (70) | 61 (39, 90) | 0.83 (0.62, 1.11) | - | 1.03 (0.76, 1.40) | |
| **Age Cohort N** | | | | **0.004** | | 0.05 |
| ≤50 years | 265 (137) | 39 (34, 48) | - | - | | |
| 51-64 years | 270 (126) | 47 (37, 66) | 0.84 (0.66, 1.07) | - | 0.84 (0.66, 1.07) | |
| ≥65 years | 276 (115) | 66 (47, 98) | **0.70 (0.54, 0.89)** | - | 0.77 (0.59, 1.00) | |
| **Age Cohort P** | | | | **0.004** | | 0.05 |
| ≤50 years | 265 (137) | 39 (34, 48) | - | - | | |
| 51-65 years | 291 (134) | 48 (37, 66) | 0.83 (0.66, 1.06) | - | 0.83 (0.65, 1.06) | |
| ≥66 years | 255 (107) | 66 (47, 98) | **0.69 (0.54, 0.89)** | - | 0.77 (0.59, 1.00) | |

[a] Number of months when 50% of patients have died/are alive, after CRS-HIPEC treatment; [b] NA upper confidence interval for an age group is due to insufficient events at later times (i.e., upper CI can't be calculated). [c] linear trend p-value; Uses R-code summary(coxph(Surv(SurvivalMonths, Dead1RtCens0) ~ OrdinalVariable, df2)), item Pr(>|z|).
[d] Adjusted for year of treatment, ASA grade, overall PCI score, number of locations with visible CPM, ever had a laparoscopy, was primary CRC tumor treated at the PMI.

Table S5. Survival median, Univariate and Multivariate Cox PH Hazard Ratios for mortality (5-year groups).

| **Age** | **N (deaths)** | **Survival median** [a] (months, 95% CI) | **Univariate Cox PH** | | **Multivariate** [d] **Cox PH** | |
|---|---|---|---|---|---|---|
| | | | **HR (95% CI)** | **p-value** | **HR (95% CI)** | **p-value** |
| **5-year groups** | | | | 0.065 [c] | | 0.057 [c] |
| <30 | 17 (12) | 32.1 (13.0, NA) [b] | - | | | |
| 30>35 | 28 (17) | 35.0 (20.0, NA) [b] | 0.61 (0.29, 1.29) | | 1.61 (0.74, 3.49) | |
| 35>40 | 59 (27) | 43.0 (34.1, NA) [b] | 0.52 (0.26, 1.03) | | 1.34 (0.65, 2.75) | |
| 40>45 | 54 (26) | 39.0 (32.0, NA) [b] | 0.56 (0.28, 1.10) | | 1.38 (0.67, 2.85) | |
| 45>50 | 84 (42) | 48.0 (30.9, NA) [b] | 0.55 (0.29, 1.04) | | 1.70 (0.85, 3.39) | |
| 50>55 | 85 (45) | 38.0 (24.0, 49.9) | 0.66 (0.35, 1.25) | | 1.72 (0.87, 3.41) | |
| 55>60 | 98 (47) | 48.0 (29.0, NA) [b] | **0.50 (0.27, 0.95)** | | 1.06 (0.55, 2.06) | |
| 60>65 | 110 (47) | 61.0 (41.0, NA) [b] | **0.44 (0.24, 0.84)** | | 1.36 (0.69, 2.68) | |
| 65>70 | 125 (45) | 95.0 (51.0, NA) [b] | **0.34 (0.18, 0.65)** | | 0.87 (0.45, 1.71) | |
| 70>75 | 88 (41) | 61.0 (35.1,106.0) | **0.48 (0.25, 0.92)** | | 1.47 (0.74, 2.93) | |
| ≥75 | 63 (29) | 56.0 (33.0, NA) [b] | **0.48 (0.24, 0.94)** | | 1.57 (0.77, 3.21) | |

[a] Survival after CRS-HIPEC treatment; [b] NA upper confidence interval for an age group is due to insufficient events at later times (i.e., upper CI can't be calculated). [c] linear trend p-value. [d] Adjusted for year of treatment, ASA grade, overall PCI score, number of locations with visible CPM, ever had a laparoscopy, was primary CRC tumor treated at the PMI

Table S6. Cox PH results for 10-year Age Cohort R.

| Age form | N (deaths) | Survival median [a] (months, 95% CI) | Univariate Cox PH for mortality | | Multivariate [c] Cox PH for mortality | |
|---|---|---|---|---|---|---|
| | | | **HR** (95% CI) | **p-value** [b] | **HR** (95% CI) | **p-value** [b] |
| **Continuous** | 811 (378) | 48 (43, 57) | 0.99 (0.98, 1.00) | **0.005** | 1.00 (0.99, 1.00) | 0.4 |
| **10-Year Bins** | | | | | | |
| **Age Cohort Q** | | | | **0.006** | | 0.54 |
| <35 yrs | 45 (29) | 34 (20, 58) | | | | |
| 35-44 yrs | 113 (53) | 41 (36, 62) | 0.73 (0.47, 1.16) | | 1.03 (0.65, 1.65) | |
| 45-54 yrs | 169 (87) | 42 (31, 54) | 0.82 (0.54, 1.26) | | 1.28 (0.82, 1.99) | |
| 55-64 yrs | 208 (94) | 54 (37, NA) | **0.64 (0.42, 0.98)** | | 0.91 (0.59, 1.39) | |
| ≥65 yrs | 276 (115) | 66 (47, 98) | **0.57 (0.38, 0.85)** | | 0.88 (0.57, 1.34) | |
| **Age Cohort R** | | | | **0.033** | | 0.2 |
| <40 yrs | 104 (56) | 37 (34, 54) | - | | - | |
| 40-49 yrs | 138 (68) | 48 (33, 58) | 0.90 (0.63, 1.29) | | 1.15 (0.80, 1.66) | |
| 50-59 yrs | 183 (92) | 41 (30, 54) | 0.93 (0.67, 1.30) | | 0.98 (0.70, 1.38) | |
| 60-69 yrs | 235 (92) | 66 (48, NA) | **0.63 (0.45, 0.88)** | | 0.80 (0.57, 1.13) | |
| ≥70 yrs | 151 (70) | 61 (39, 90) | 0.78 (0.55, 1.12) | | 1.11 (0.77, 1.61) | |

[a] Number of months when 50% of patients have died/are alive, after CRS-HIPEC treatment. [b] linear trend p-value for ternary/quaternary/10-year Age Cohorts.
[c] Adjusted for year of treatment, ASA grade, overall PCI score, number of locations with visible CPM, ever had a laparoscopy, was primary CRC tumor treated at the PMI

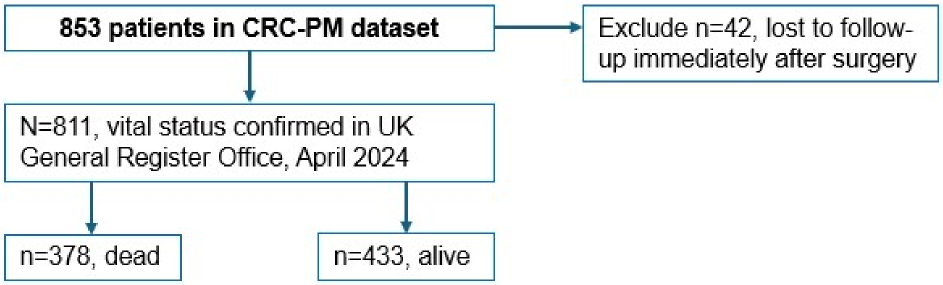

Figure S1. Patient selection process for the study.

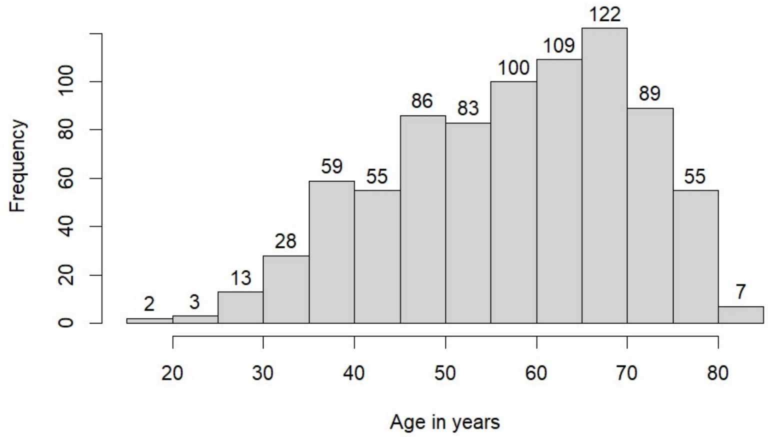

Figure S2. Age distribution of 811 patients included in the study.

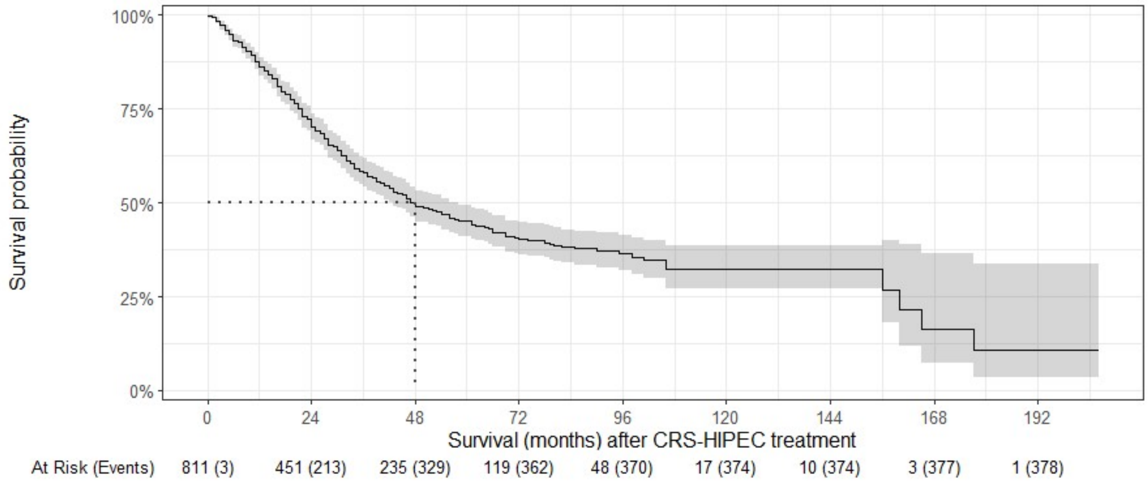

Figure S3. K-M plot, 811 CPM patients included in the study; shows survival median value and 95% CI band.

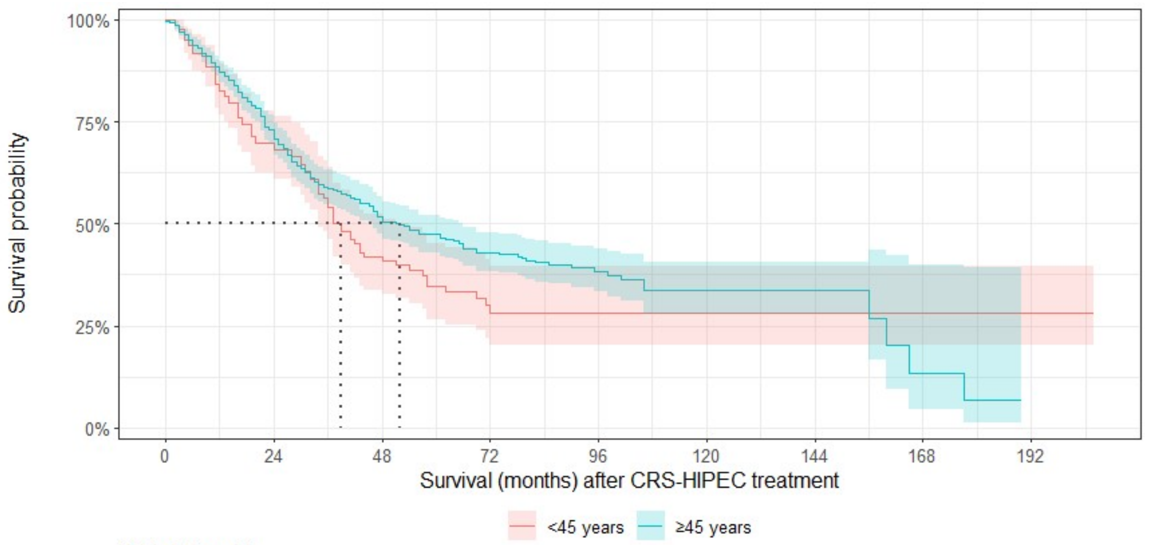

At Risk (Events)

| | | | | | | | | | |
|---|---|---|---|---|---|---|---|---|---|
| <45 years | 158 (0) | 88 (45) | 40 (73) | 16 (82) | 5 (82) | 1 (82) | 1 (82) | 1 (82) | 1 (82) |
| ≥45 years | 653 (3) | 363 (168) | 195 (256) | 103 (280) | 43 (288) | 16 (292) | 9 (292) | 2 (295) | 0 (296) |

Figure S4. K-M plot <45 vs. ≥45 years (current US CRC screening age cut-off); shows survival medians and 95% CI bands.

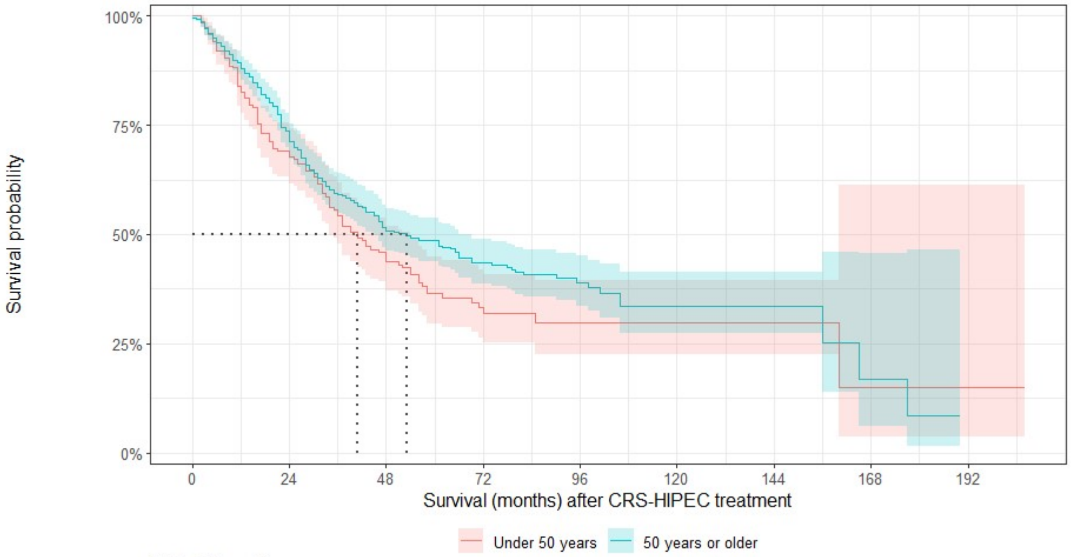

At Risk (Events)

| | | | | | | | | | |
|---|---|---|---|---|---|---|---|---|---|
| Under 50 years | 242 (0) | 135 (70) | 67 (109) | 27 (122) | 9 (123) | 4 (123) | 4 (123) | 1 (124) | 1 (124) |
| 50 years or older | 569 (3) | 316 (143) | 168 (220) | 92 (240) | 39 (247) | 13 (251) | 6 (251) | 2 (253) | 0 (254) |

Figure S5. K-M plot <50 vs. ≥50 years (Age Cohort B; (prior US CRC screening age cut-off); shows survival median values and 95% CI bands.

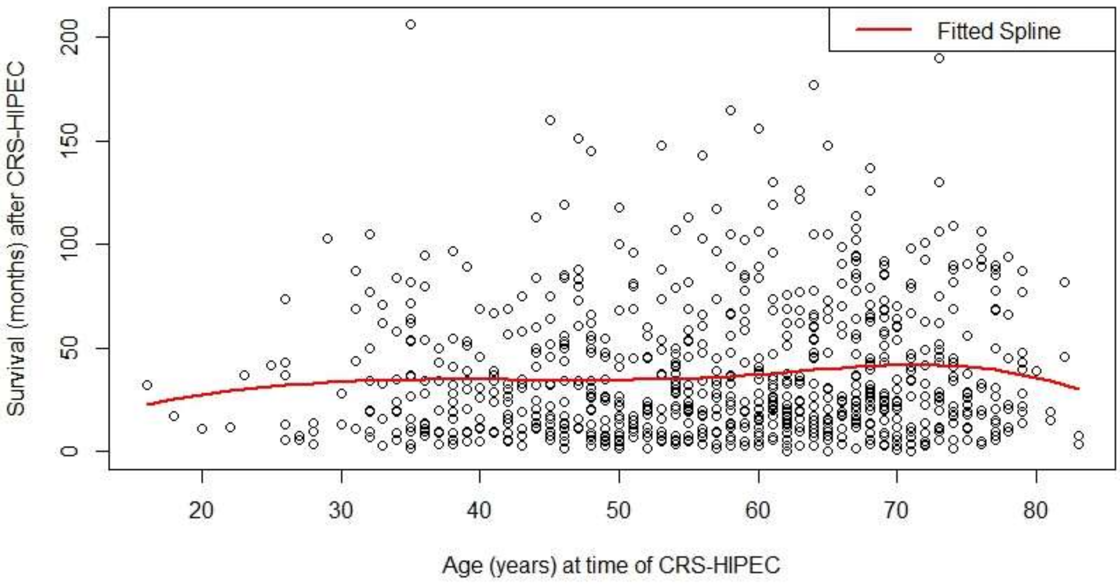

Figure S6. B-spline regression (4 degrees freedom) for survival by age at time of CRS-HIPEC.

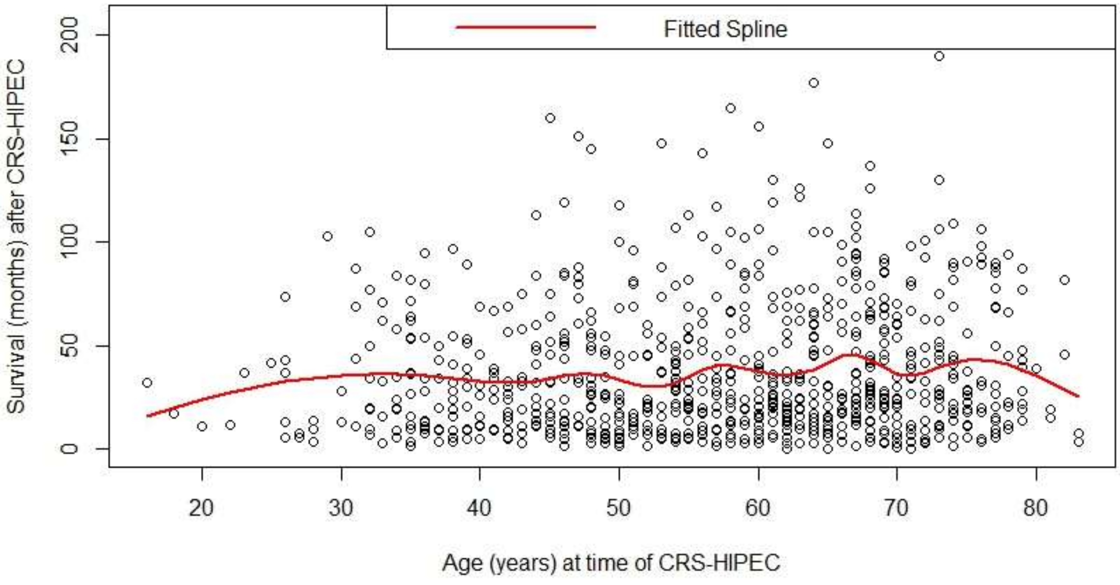

Figure S7. B-spline regression (13 degrees freedom) for survival by age at time of CRS-HIPEC.

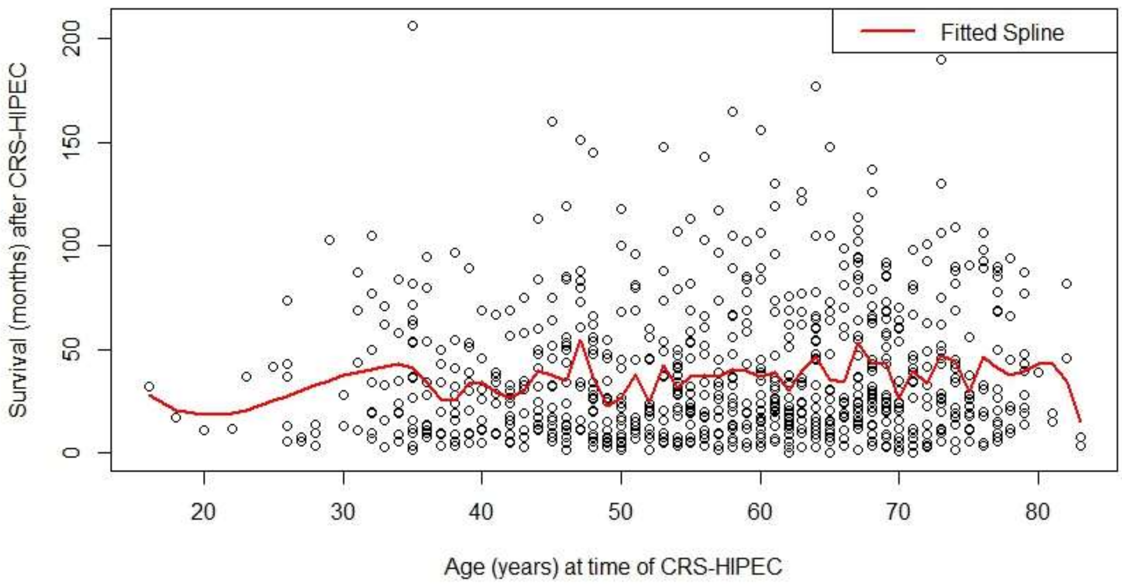

Figure S8. B-spline regression (50 degrees freedom) for survival by age at time of CRS-HIPEC.

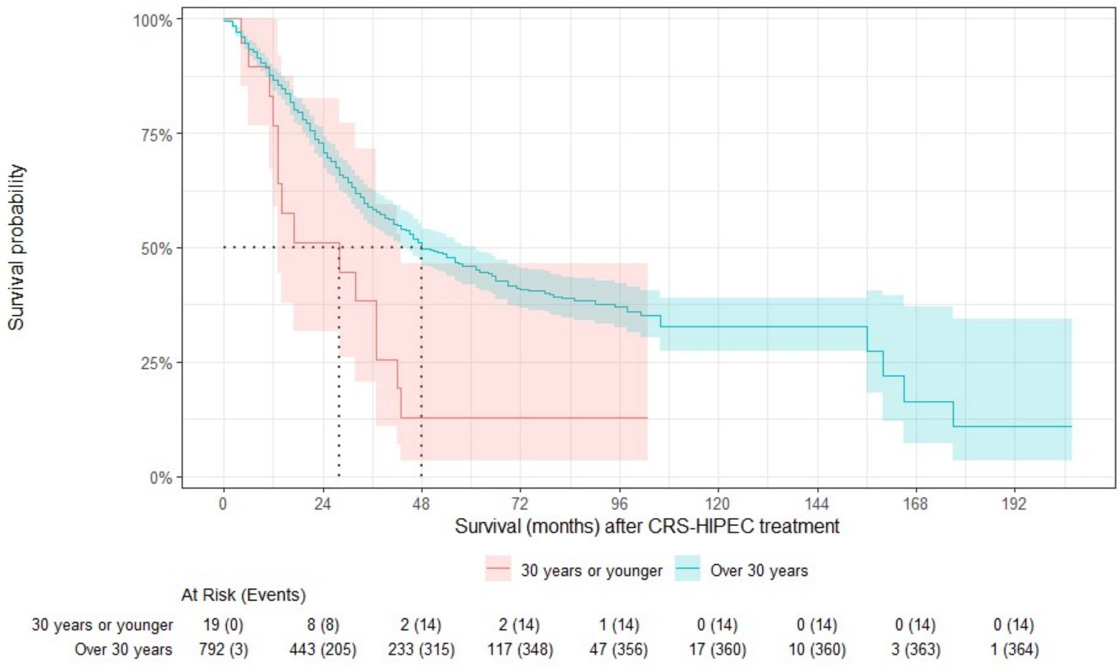

Figure S9. K-M plot ≤30 vs. >30 years; shows survival medians and 95% CI bands.

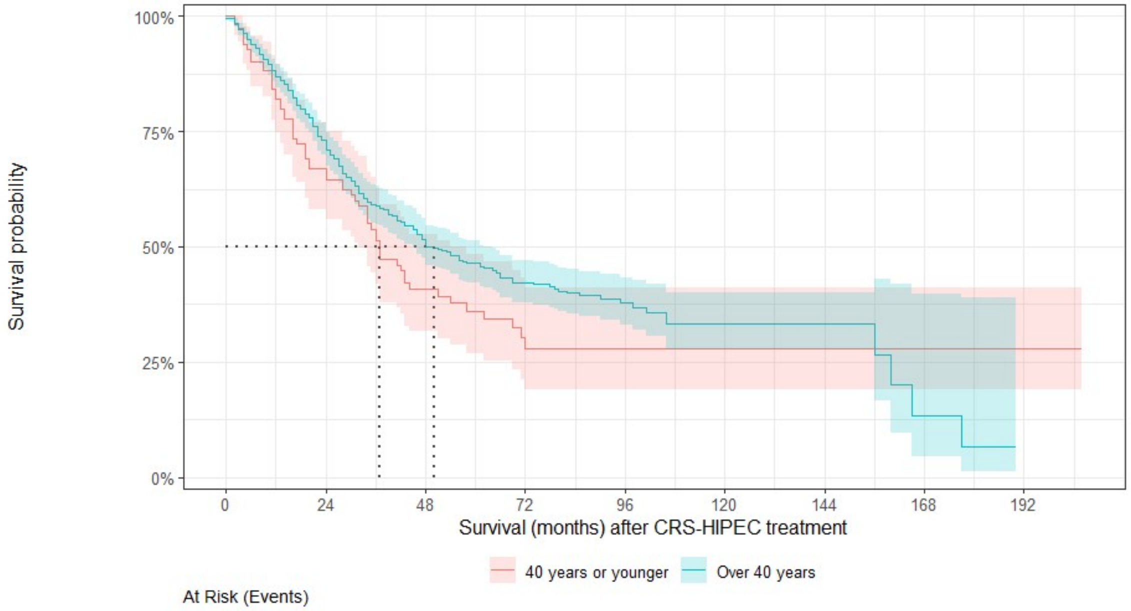

At Risk (Events)

| | | | | | | | | | |
|---|---|---|---|---|---|---|---|---|---|
| 40 years or younger | 113 (0) | 60 (35) | 30 (54) | 13 (61) | 4 (61) | 1 (61) | 1 (61) | 1 (61) | 1 (61) |
| Over 40 years | 698 (3) | 391 (178) | 205 (275) | 106 (301) | 44 (309) | 16 (313) | 9 (313) | 2 (316) | 0 (317) |

Figure S10. K-M plot ≤40 vs. >40 years (Age Cohort A); shows survival medians and 95% CI bands.

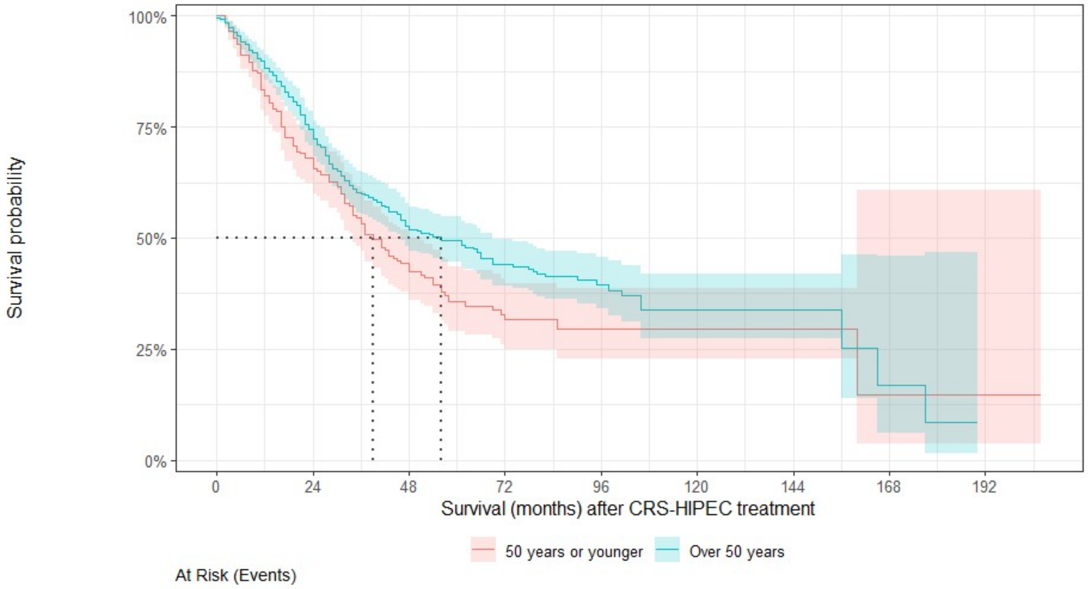

At Risk (Events)

| | | | | | | | | | |
|---|---|---|---|---|---|---|---|---|---|
| 50 years or younger | 265 (0) | 144 (81) | 70 (122) | 29 (135) | 11 (136) | 4 (136) | 4 (136) | 1 (137) | 1 (137) |
| Over 50 years | 546 (3) | 307 (132) | 165 (207) | 90 (227) | 37 (234) | 13 (238) | 6 (238) | 2 (240) | 0 (241) |

Figure S11. K-M plot ≤50 vs. >50 years (Age Cohort C); shows survival medians and 95% CI bands

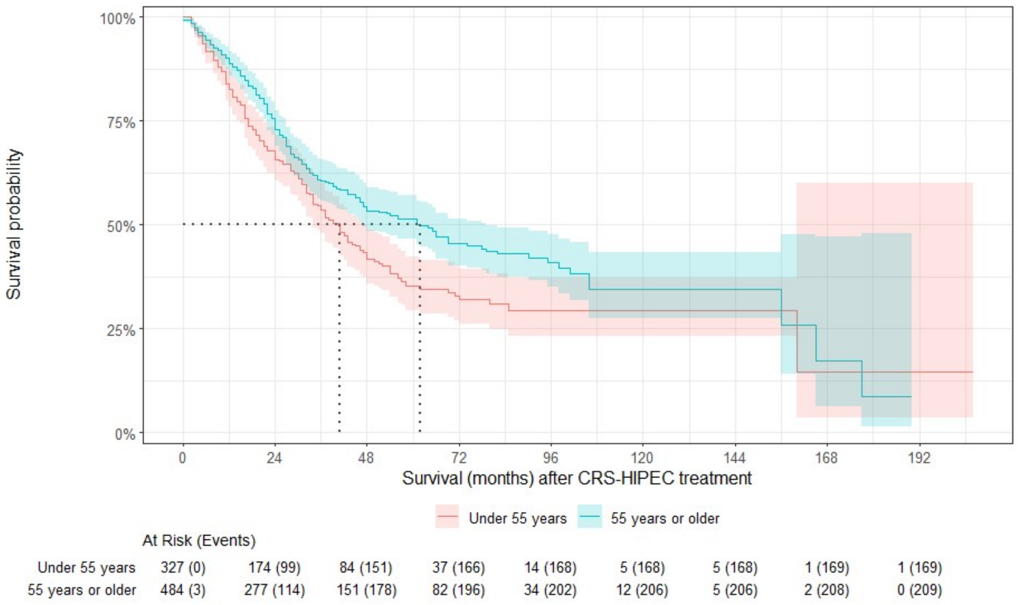

Figure S12. K-M plot <55 vs. ≥55 years (Age Cohort D); shows survival medians and 95% CI bands.

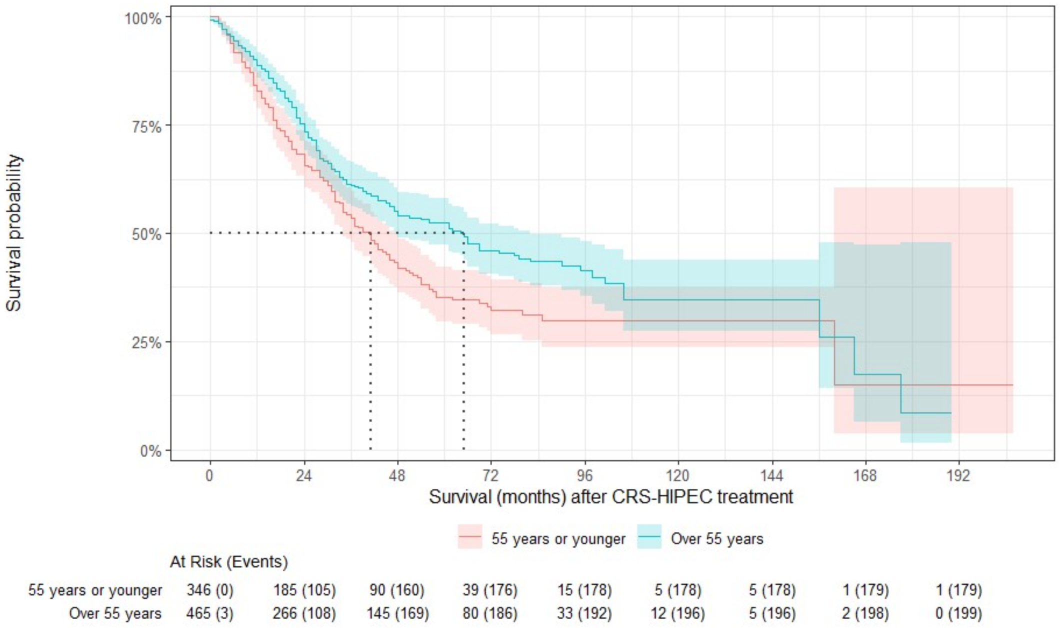

Figure S13. K-M plot ≤55 vs. >55 years (Age Cohort E); shows survival medians and 95% CI bands.

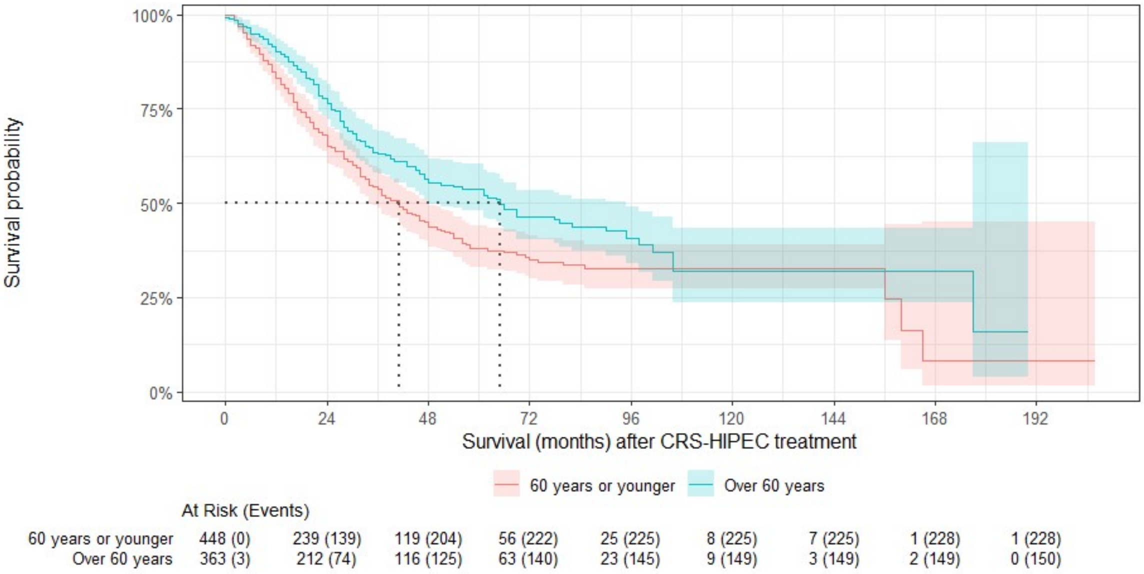

Figure S14. K-M plot, Age Cohort G (≤60 vs. >60 years); shows survival medians and 95% CI bands.

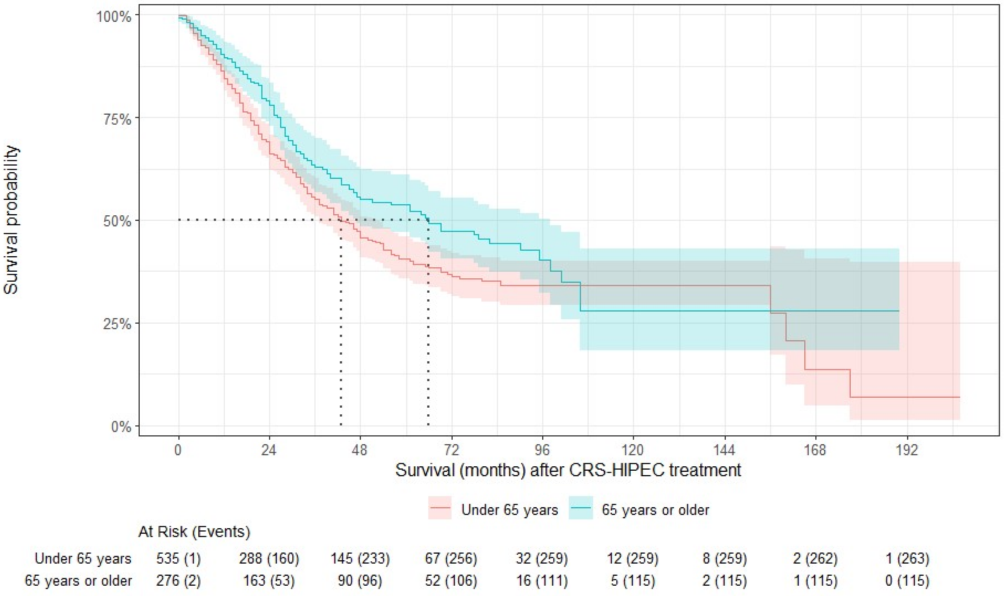

Figure S15. K-M plot <65 vs. ≥65 years (Age Cohort H); shows survival medians and 95% CI bands.

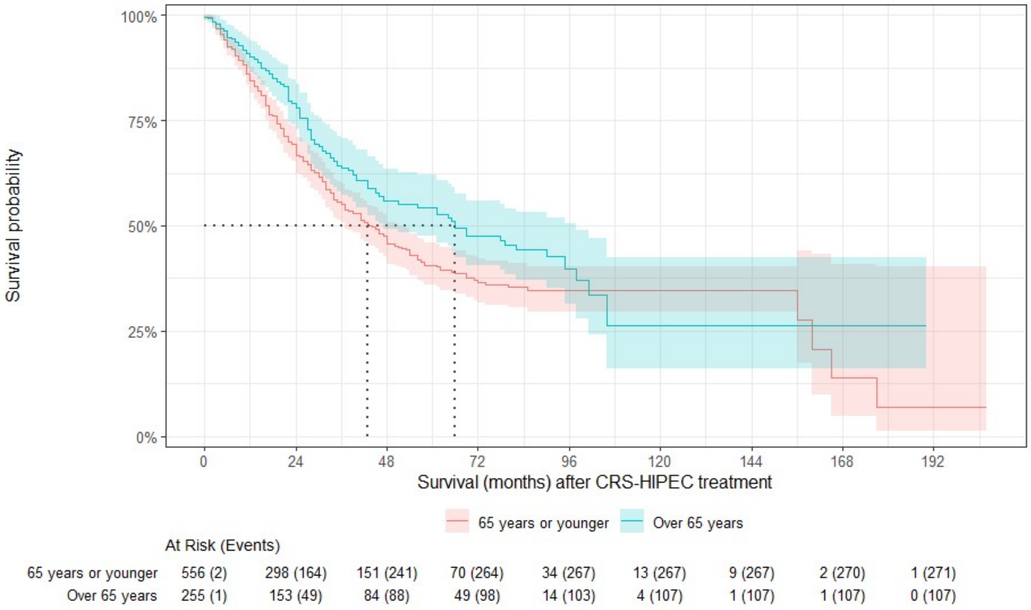

Figure S16. K-M plot ≤65 vs. >65 years (Age Cohort J); shows survival medians and 95% CI bands.

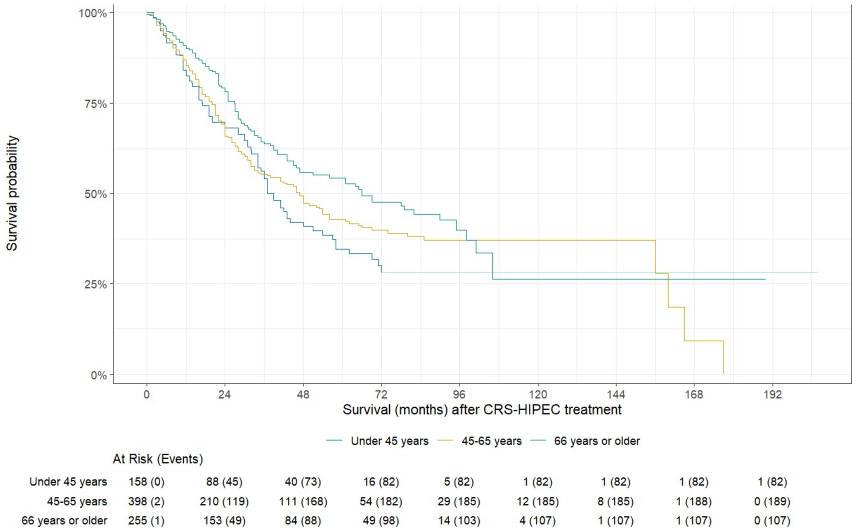

Figure S17. K-M plot <45 vs. 45-65 vs. ≥66 years (Age Cohort K).

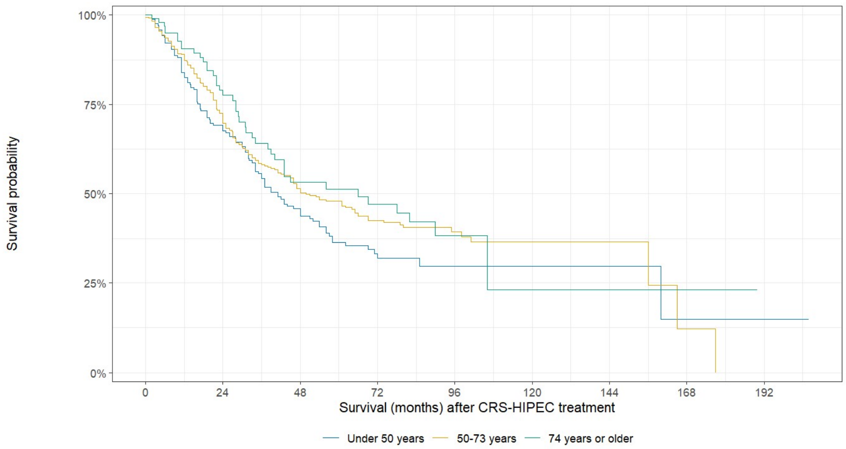

At Risk (Events)

| | | | | | | | | | |
|---|---|---|---|---|---|---|---|---|---|
| Under 50 years | 242 (0) | 135 (70) | 67 (109) | 27 (122) | 9 (123) | 4 (123) | 4 (123) | 1 (124) | 1 (124) |
| 50-73 years | 470 (3) | 260 (124) | 137 (185) | 71 (202) | 33 (206) | 11 (208) | 5 (208) | 1 (210) | 0 (211) |
| 74 years or older | 99 (0) | 56 (19) | 31 (35) | 21 (38) | 6 (41) | 2 (43) | 1 (43) | 1 (43) | 0 (43) |

Figure S18. K-M plot <50 vs. 50-73 vs. ≥74 years (Age Cohort L).

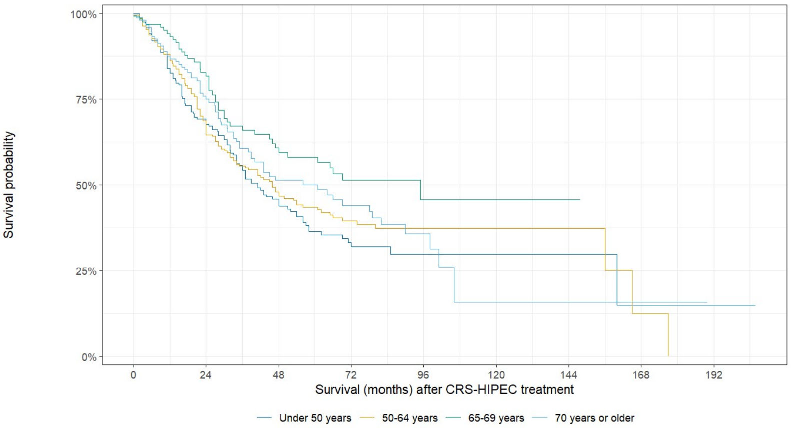

At Risk (Events)

| | | | | | | | | | |
|---|---|---|---|---|---|---|---|---|---|
| Under 50 years | 242 (0) | 135 (70) | 67 (109) | 27 (122) | 9 (123) | 4 (123) | 4 (123) | 1 (124) | 1 (124) |
| 50-64 years | 293 (1) | 153 (90) | 78 (124) | 40 (134) | 23 (136) | 8 (136) | 4 (136) | 1 (138) | 0 (139) |
| 65-69 years | 125 (1) | 79 (20) | 46 (39) | 26 (44) | 8 (45) | 3 (45) | 1 (45) | 0 (45) | 0 (45) |
| 70 years or older | 151 (1) | 84 (33) | 44 (57) | 26 (62) | 8 (66) | 2 (70) | 1 (70) | 1 (70) | 0 (70) |

Figure S19. K-M plot <45 vs. 50-64 vs. 65-69 vs. ≥70 years (Age Cohort M).

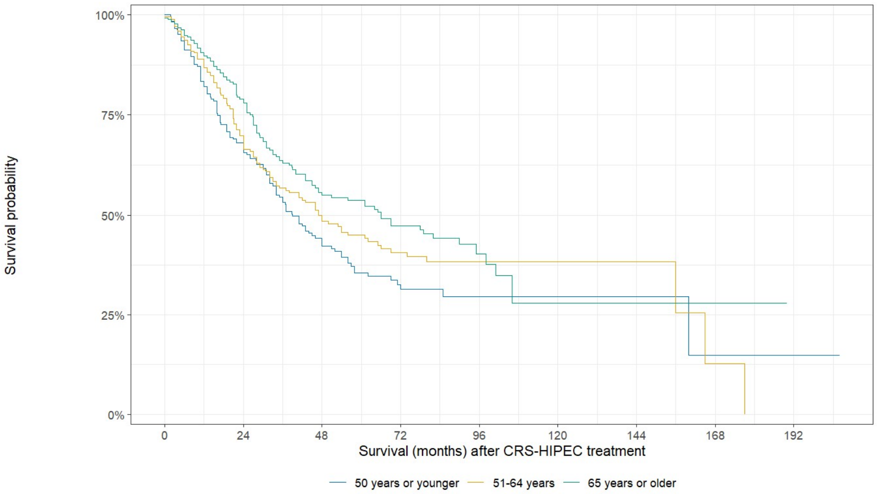

At Risk (Events)

| | 0 | 24 | 48 | 72 | 96 | 120 | 144 | 168 | 192 |
|---|---|---|---|---|---|---|---|---|---|
| 50 years or younger | 265 (0) | 144 (81) | 70 (122) | 29 (135) | 11 (136) | 4 (136) | 4 (136) | 1 (137) | 1 (137) |
| 51-64 years | 270 (1) | 144 (79) | 75 (111) | 38 (121) | 21 (123) | 8 (123) | 4 (123) | 1 (125) | 0 (126) |
| 65 years or older | 276 (2) | 163 (53) | 90 (96) | 52 (106) | 16 (111) | 5 (115) | 2 (115) | 1 (115) | 0 (115) |

Figure S20. K-M plot <50 vs. 51-64 vs. ≥65 years (Age Cohort N).

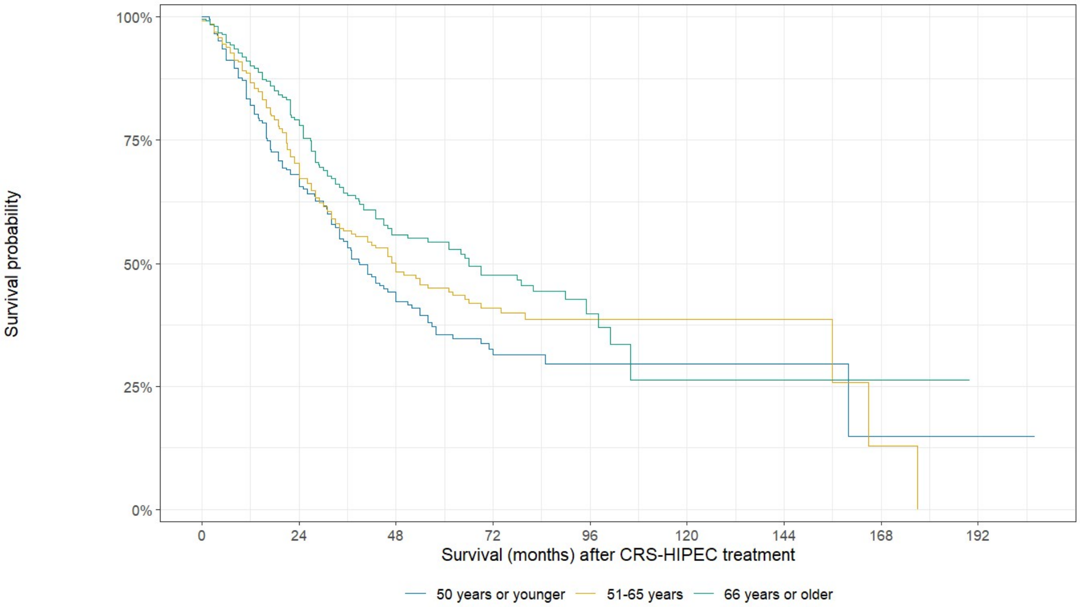

At Risk (Events)

| | 0 | 24 | 48 | 72 | 96 | 120 | 144 | 168 | 192 |
|---|---|---|---|---|---|---|---|---|---|
| 50 years or younger | 265 (0) | 144 (81) | 70 (122) | 29 (135) | 11 (136) | 4 (136) | 4 (136) | 1 (137) | 1 (137) |
| 51-65 years | 291 (2) | 154 (83) | 81 (119) | 41 (129) | 23 (131) | 9 (131) | 5 (131) | 1 (133) | 0 (134) |
| 66 years or older | 255 (1) | 153 (49) | 84 (88) | 49 (98) | 14 (103) | 4 (107) | 1 (107) | 1 (107) | 0 (107) |

Figure S21. K-M plot ≤50 vs. 51-65 vs. ≥66 years (Age Cohort P)..

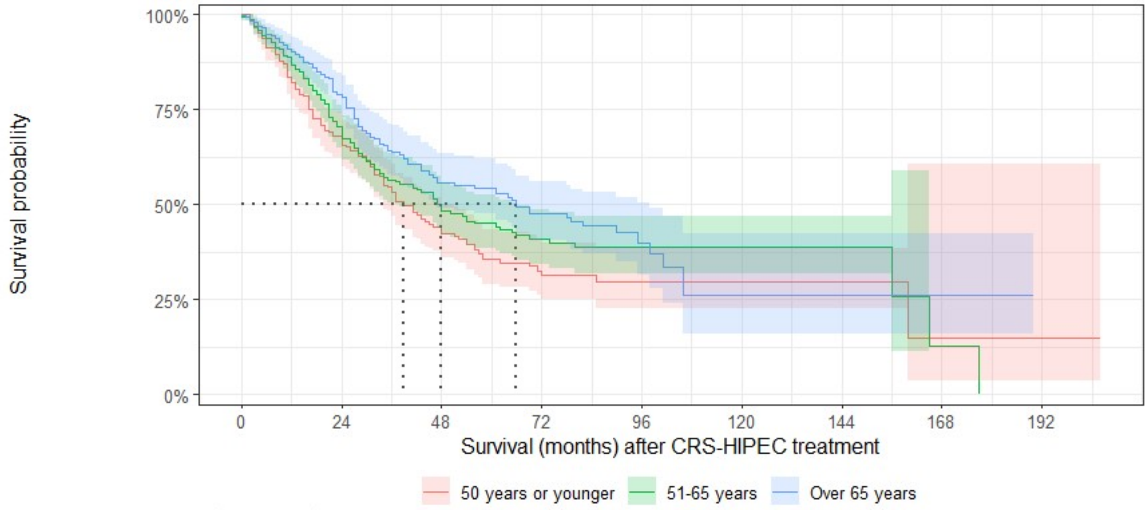

At Risk (Events)

| | | | | | | | | | |
|---|---|---|---|---|---|---|---|---|---|
| 50 years or younger | 265 (0) | 144 (81) | 70 (122) | 29 (135) | 11 (136) | 4 (136) | 4 (136) | 1 (137) | 1 (137) |
| 51-65 years | 291 (2) | 154 (83) | 81 (119) | 41 (129) | 23 (131) | 9 (131) | 5 (131) | 1 (133) | 0 (134) |
| Over 65 years | 255 (1) | 153 (49) | 84 (88) | 49 (98) | 14 (103) | 4 (107) | 1 (107) | 1 (107) | 0 (107) |

Figure S22. K-M plot, Age Cohort P (≤50 vs. 51-65 vs. ≥66 years); shows survival medians and 95% CI bands

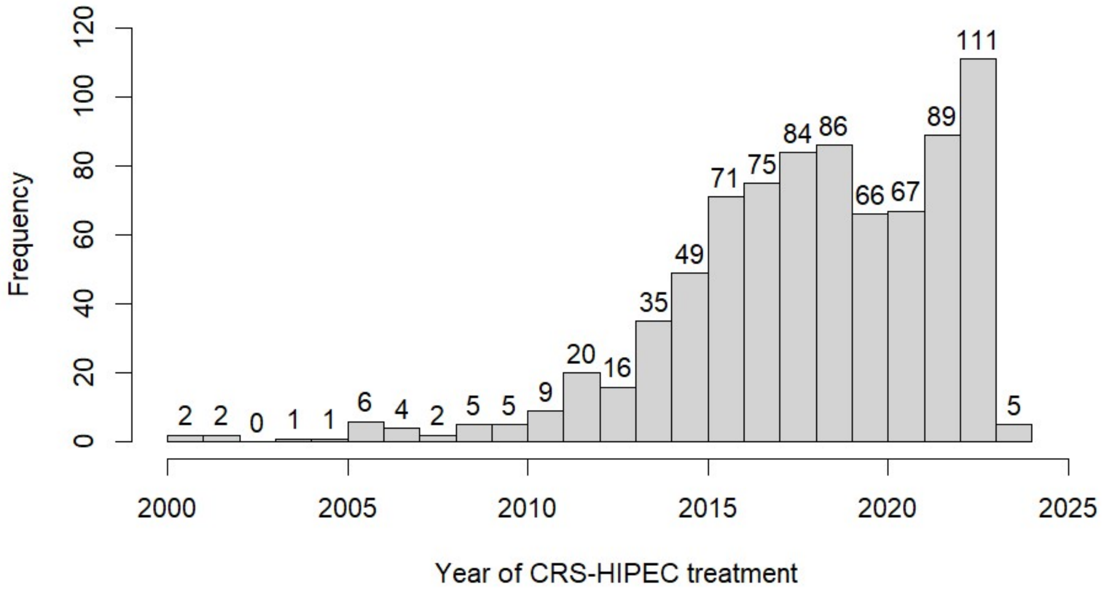

Figure S23. Distribution of CRS-HIPEC treatment year for the study's 811 patients.

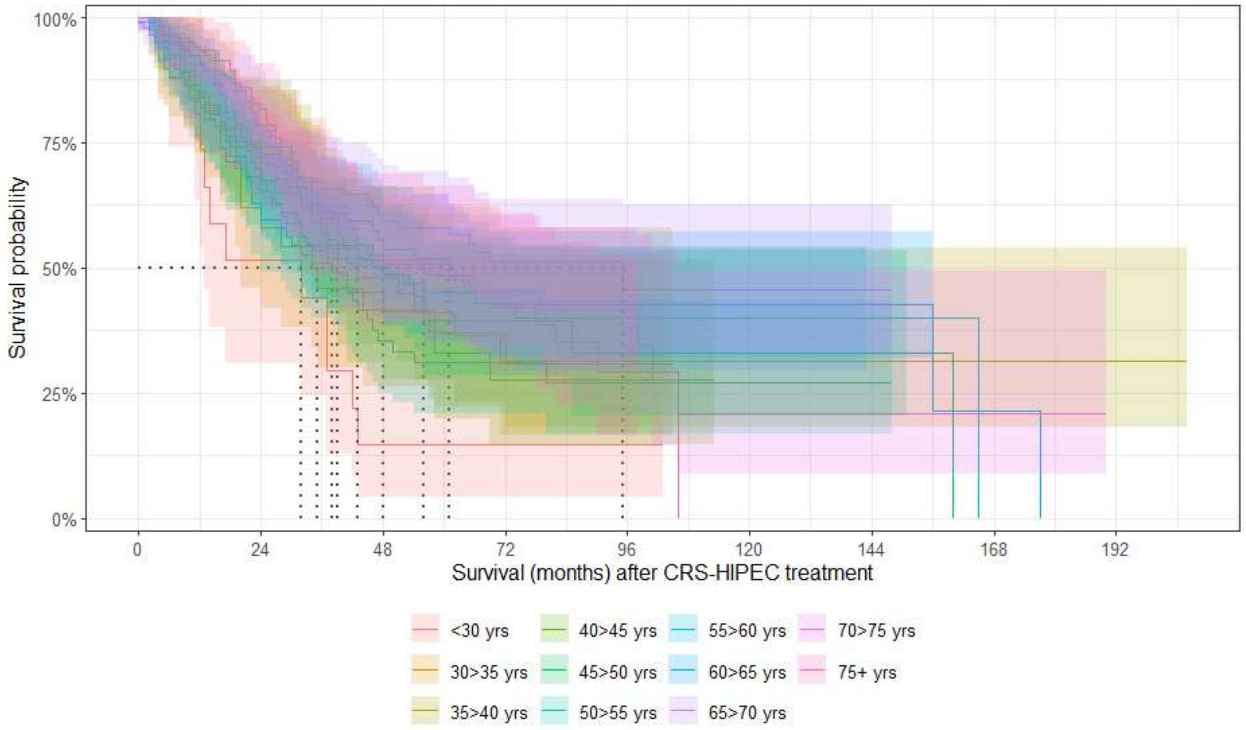

At Risk (Events)

| | | | | | | | | | |
|---|---|---|---|---|---|---|---|---|---|
| <30 yrs | 17 (0) | 7 (7) | 2 (12) | 2 (12) | 1 (12) | 0 (12) | 0 (12) | 0 (12) | 0 (12) |
| 30>35 yrs | 28 (0) | 16 (11) | 10 (15) | 4 (17) | 1 (17) | 0 (17) | 0 (17) | 0 (17) | 0 (17) |
| 35>40 yrs | 59 (0) | 32 (14) | 16 (23) | 7 (27) | 2 (27) | 1 (27) | 1 (27) | 1 (27) | 1 (27) |
| 40>45 yrs | 54 (0) | 33 (13) | 12 (23) | 3 (26) | 1 (26) | 0 (26) | 0 (26) | 0 (26) | 0 (26) |
| 45>50 yrs | 84 (0) | 47 (25) | 27 (36) | 11 (40) | 4 (41) | 3 (41) | 3 (41) | 0 (42) | 0 (42) |
| 50>55 yrs | 85 (0) | 39 (29) | 17 (42) | 10 (44) | 5 (45) | 1 (45) | 1 (45) | 0 (45) | 0 (45) |
| 55>60 yrs | 98 (0) | 54 (30) | 31 (42) | 15 (45) | 9 (46) | 2 (46) | 1 (46) | 0 (47) | 0 (47) |
| 60>65 yrs | 110 (1) | 60 (31) | 30 (40) | 15 (45) | 9 (45) | 5 (45) | 2 (45) | 1 (46) | 0 (47) |
| 65>70 yrs | 125 (1) | 79 (20) | 46 (39) | 26 (44) | 8 (45) | 3 (45) | 1 (45) | 0 (45) | 0 (45) |
| 70>75 yrs | 88 (1) | 51 (22) | 25 (35) | 13 (37) | 6 (38) | 2 (41) | 1 (41) | 1 (41) | 0 (41) |
| 75+ yrs | 63 (0) | 33 (11) | 19 (22) | 13 (25) | 2 (28) | 0 (29) | 0 (29) | 0 (29) | 0 (29) |

Figure S24. K-M plot, 5-year age bins; shows survival medians and 95% CI bands.

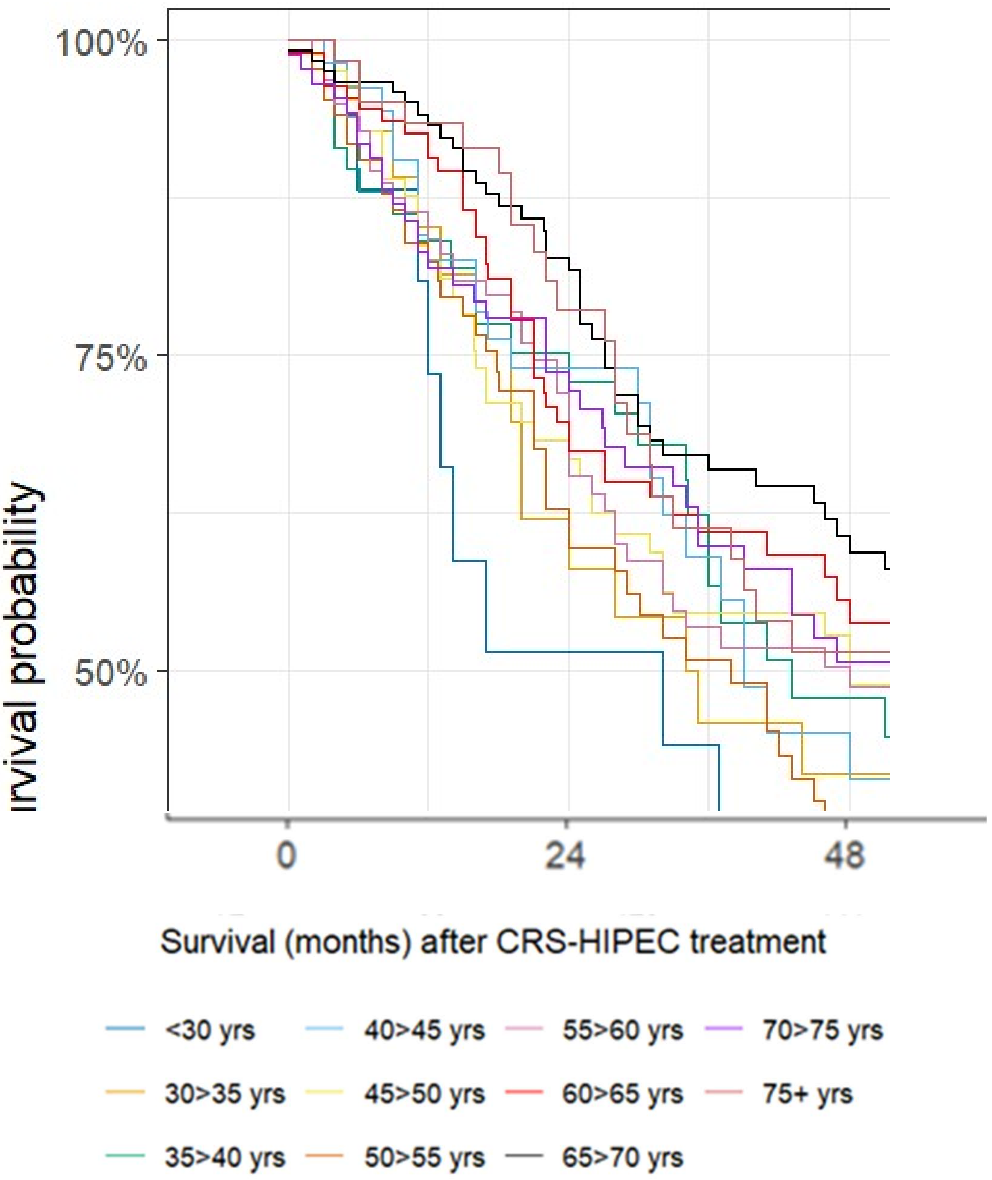

Figure S25. Overlap detail in K-M plot, first 48 months for patients in 5-year age bins. *NOTE: This is three portions of Figure S24, magnified and cropped (curve, x-axis tick legend, and X-axis title with line color legend*

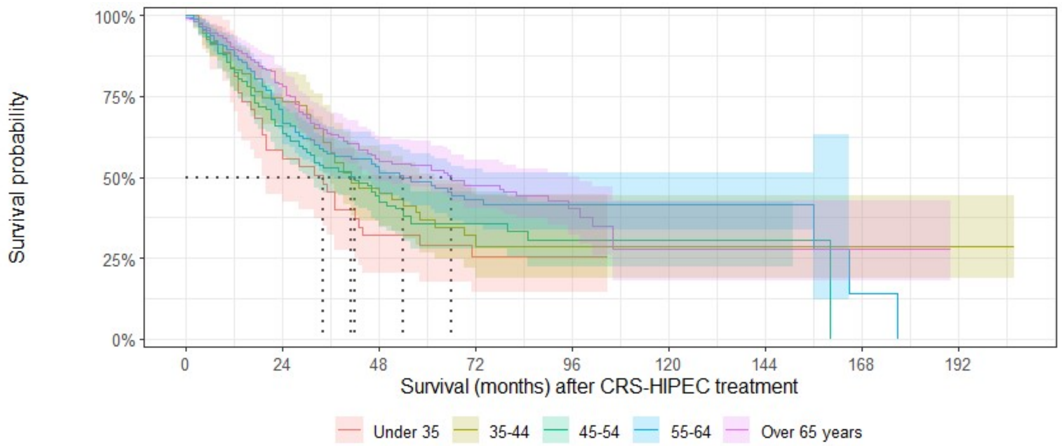

At Risk (Events)

| | | | | | | | | | |
|---|---|---|---|---|---|---|---|---|---|
| Under 35 | 45 (0) | 23 (18) | 12 (27) | 6 (29) | 2 (29) | 0 (29) | 0 (29) | 0 (29) | 0 (29) |
| 35-44 | 113 (0) | 65 (27) | 28 (46) | 10 (53) | 3 (53) | 1 (53) | 1 (53) | 1 (53) | 1 (53) |
| 45-54 | 169 (0) | 86 (54) | 44 (78) | 21 (84) | 9 (86) | 4 (86) | 4 (86) | 0 (87) | 0 (87) |
| 55-64 | 208 (1) | 114 (61) | 61 (82) | 30 (90) | 18 (91) | 7 (91) | 3 (91) | 1 (93) | 0 (94) |
| Over 65 years | 276 (2) | 163 (53) | 90 (96) | 52 (106) | 16 (111) | 5 (115) | 2 (115) | 1 (115) | 0 (115) |

Figure S26. K-M plot, 10-year age groups (starting with <35 years, Age Cohort Q); shows survival medians and 95% CI bands.

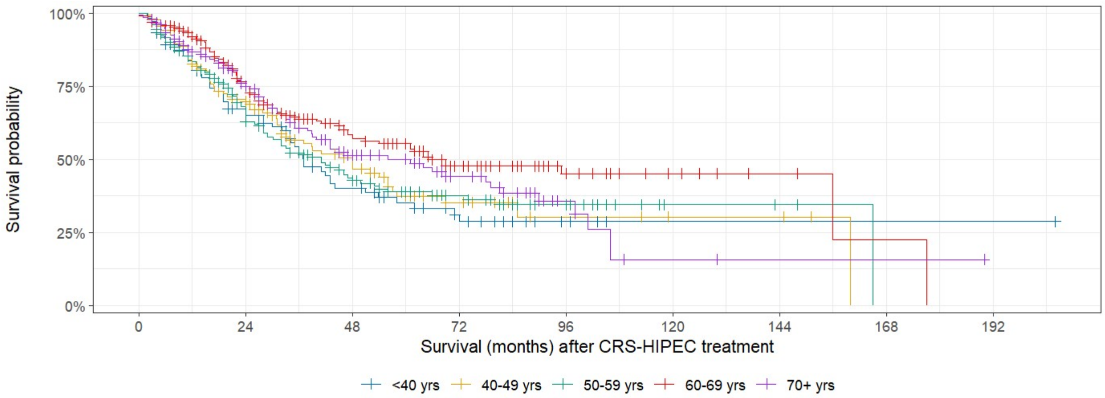

At Risk (Events)

| | | | | | | | | | |
|---|---|---|---|---|---|---|---|---|---|
| <40 yrs | 104 (0) | 55 (32) | 28 (50) | 13 (56) | 4 (56) | 1 (56) | 1 (56) | 1 (56) | 1 (56) |
| 40-49 yrs | 138 (0) | 80 (38) | 39 (59) | 14 (66) | 5 (67) | 3 (67) | 3 (67) | 0 (68) | 0 (68) |
| 50-59 yrs | 183 (0) | 93 (59) | 48 (84) | 25 (89) | 14 (91) | 3 (91) | 2 (91) | 0 (92) | 0 (92) |
| 60-69 yrs | 235 (2) | 139 (51) | 76 (79) | 41 (89) | 17 (90) | 8 (90) | 3 (90) | 1 (91) | 0 (92) |
| 70+ yrs | 151 (1) | 84 (33) | 44 (57) | 26 (62) | 8 (66) | 2 (70) | 1 (70) | 1 (70) | 0 (70) |

Figure S27. K-M plot, 10-year age groups (starting with <40 years, Age Cohort R).

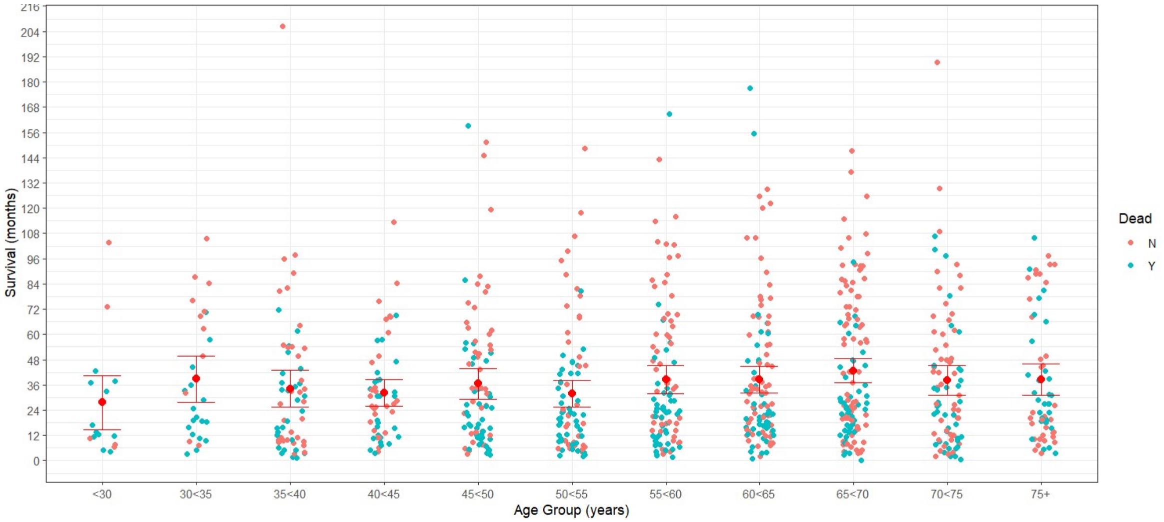

Figure S28. Months of post-treatment follow-up and survival for CPM patients by 5-year age group (age at time of CRS-HIPEC; mean, 95% CI).

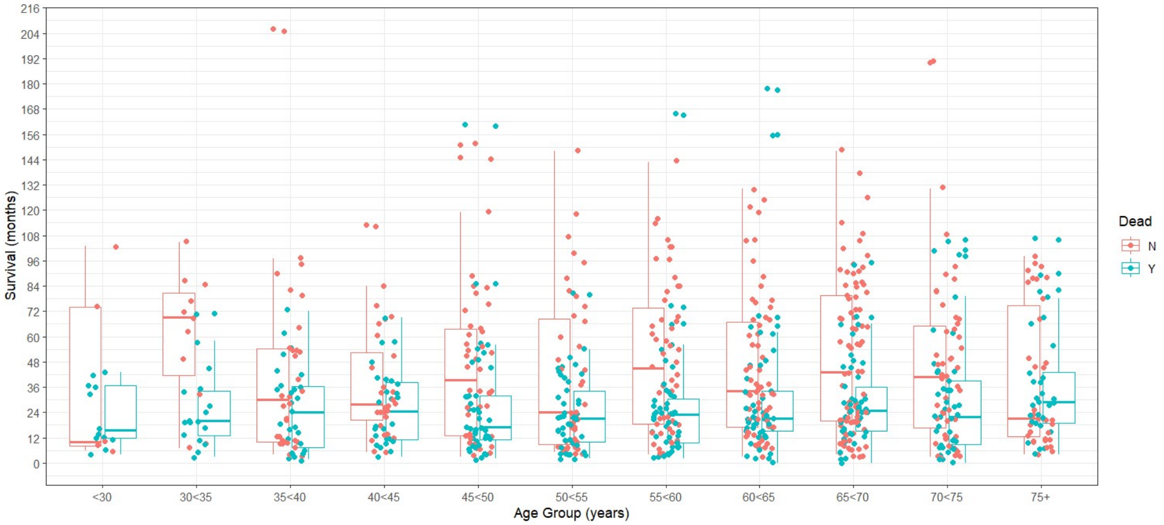

Figure S29. Months of post-treatment follow-up and survival for CPM patients by 5-year age group (age at time of CRS-HIPEC; mean, boxplots by vital status)
